# Using low-cost sensors and GPS to assess spatiotemporal variations in personal exposure to PM_2.5_ in the Washington State Twin Registry

**DOI:** 10.1101/2025.06.09.25329147

**Authors:** Ningrui Liu, Ally Avery, Elena Austin, John S. Meschke, Nicola K. Beck, Graeme Carvlin, Yisi Liu, Anne V. Moudon, Igor Novosselov, Jeffry H. Shirai, Glen E. Duncan, Edmund Seto

## Abstract

Epidemiological studies typically rely on exposure assessments based on ambient PM_2.5_ concentrations at participants’ home addresses. However, these approaches neglect personal exposures indoors and across different non-residential microenvironments. To address this problem, our study combined low-cost sensors and GPS to conduct two-week personal PM_2.5_ monitoring in 168 adults recruited from the Washington State Twin Registry between 2018 and 2021. PM_2.5_ mass concentration, size-resolved particle number concentration, temperature, humidity, and GPS coordinates were recorded at 1-minute intervals, providing 5,161,737 datapoints. We used GPS coordinates and a processing algorithm for automatic classification of microenvironments, including seven land use types and vehicles, and time spent indoors/outdoors. The low-cost sensors were calibrated in-situ, using regulatory monitoring data within 600 m of participants’ outdoor measurements (R^2^ = 0.93). A linear mixed model was used to estimate the associations of multiple spatiotemporal factors with personal exposure concentrations. The average PM_2.5_ exposure concentration was 8.1 ± 15.8 μg/m^3^ for all participants. Indoor exposure concentration was higher than outdoor exposure level, and indoor exposure dose contributed 77% to the total exposure. Exposures in residential and industrial land use had a higher concentration than in other areas, and accounted for 69% of the total exposure dose. Furthermore, personal exposure concentration was the highest during winter and evening hours, possibly due to cooking and heating-related behaviors. This study demonstrates that personal monitoring can capture spatiotemporal variations in PM_2.5_ exposure more accurately than home-based approaches based on ambient air quality, and suggests opportunities for controlling exposures in certain microenvironments.

TOC Art

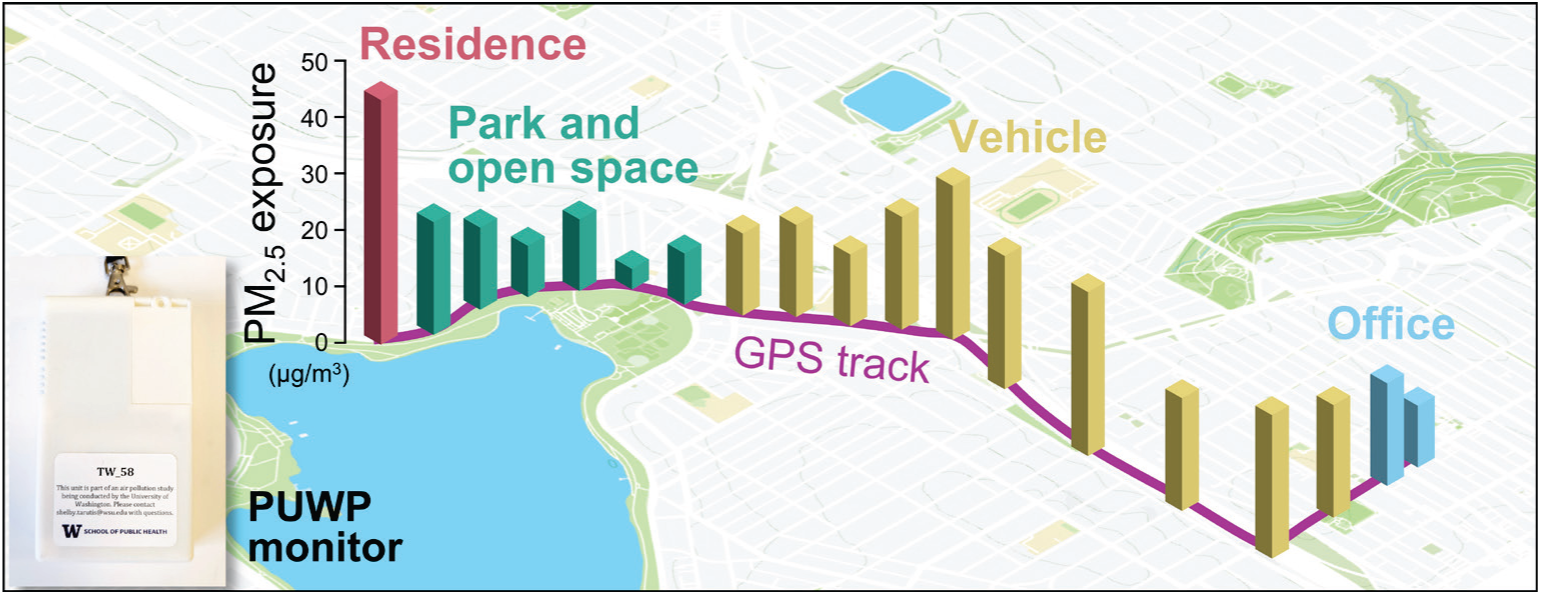

**Highlights:**

● A total of 168 participants completed two-week personal PM_2.5_ and GPS monitoring.
● Personal exposure to PM_2.5_ had substantial spatiotemporal variation.
● Indoor exposure had higher exposure concentration and exposure dose than outdoor.
● Residential/industrial PM_2.5_ concentration was higher based on regression analysis.
● Home-based exposure assessment cannot capture actual personal exposure patterns.

## 1 Introduction

Fine particulate matter (PM_2.5_) is associated with multiple adverse health outcomes, such as lung cancer and cardiovascular diseases. Despite concerted emission regulation and public health efforts, air pollution and especially PM_2.5_ levels remain a challenging problem in many countries. The Global Burden of Diseases, Injuries, and Risk Factors Study (GBD) has documented that PM_2.5_ was one of the top risk factors globally in 2021, contributing 8.0% of the total disability-adjusted life years (DALYs) ^1^. It is therefore essential to accurately assess the individual– and population-level exposure to PM_2.5_ to quantify health impacts.

Large epidemiological studies typically rely on geospatial models and data from ambient air quality monitoring stations or remote sensing to obtain the ambient PM_2.5_ concentrations at/near participants’ home addresses, which are then linked to health outcomes ^2–8^. The possible reasons are that ambient air pollution is commonly regulated in many countries, and such studies can inform policy development. It is also relatively difficult to measure personal exposure. However, these approaches include two assumptions. The first assumption is that outdoor exposure is an unbiased representation of their total exposure which includes both indoor and outdoor exposure. As people spend over 80% of their lifetime in indoor environments ^9, 10^, there is important bias introduced by this assumption because of variability in infiltration rates and generation of PM_2.5_ indoors from sources such as cooking, heating, smoking, and cleaning ^11–14^. The large variation in indoor PM_2.5_ levels can be related to household characteristics, residents’ behavioral characteristics, socio-economic status, and local climate ^15–18^. The second assumption is that people spend most of their time at home, so that estimating exposure at the home address results in the best and least biased estimate of total exposure. This assumption fails to account for important time-activity patterns where people usually move and stay in multiple non-residential microenvironments in a day, such as working in an office, having meals in restaurants, and commuting in vehicles. It also fails to account for the important relationship between time-activity patterns and demographic characteristics, including age, gender, employment status, and housing type ^19^. The indoor PM_2.5_ concentrations in different microenvironments can also vary substantially and be quite different from concentrations in homes ^20–23^. The above evidence suggests that home-based modeling methods may not accurately represent the personal exposures to PM_2.5_ people experience on a daily basis under “real-world” conditions. Some previous studies also found very weak correlations between personal exposure and outdoor PM_2.5_ levels measured at fixed sites ^24–29^, further complicating the use of traditional exposure methods in exposure-health studies.

To address the extent of what is noted above and related problems, researchers have started to equip subjects with newer, low-cost portable/wearable sensors or samplers to obtain personal exposure data^20, 21, 24, 30–49^. A few of these studies used the active sampler to collect filter samples of PM_2.5_ across one or more weeks and obtain average exposures over the monitoring period, which were further utilized for component analysis and source apportionment ^35, 43, 46^. Most other studies leveraged low-cost sensors to monitor real-time exposures and apportioned the personal exposure of PM_2.5_ into various microenvironments, such as home, workplace, transit, restaurant, and school, to capture the spatiotemporal variability of exposure levels ^20, 21, 24, 30, 31, 33, 34, 37–39,42, 45, 48^.

However, these approaches still have limitations. From the perspective of identifying microenvironments, some studies relied on time-activity diaries and questionnaires, which is burdensome for relatively long time periods ^31, 32, 34, 35, 37, 39, 41^. The subjective nature of these data may also lead to recall bias, which may misclassify some visited microenvironments and may not record the exposure time in different microenvironments accurately ^50, 51^. Some studies leveraged the Global Positioning System (GPS) receiver to track subject’s movement, but then manually identified the microenvironments on a map according to the GPS coordinates and validated them with time-activity diaries, if available ^20, 21, 38^. Microenvironments often included home, workplace, and school, while other microenvironments, such as parks and restaurants, were usually neglected ^30, 31, 33, 34, 38, 39, 48^. Further, personal exposure in workplaces was too broad, ignoring that the workplaces can cover a variety of microenvironments, such as offices, factories, and restaurants, where the PM_2.5_ concentrations can vary substantially. From a temporal perspective, the personal exposure monitoring usually lasted for a relatively short period, i.e., 1 to 2 days ^24, 30, 31, 34, 36, 39, 41, 43, 45^, with very few covering more than two weeks ^35, 37, 47^. Using short monitoring periods is likely due to the high cost of equipment, burden on participants, time, and labor. Additionally, the calibration of these low-cost sensors is an important issue. Many studies on personal exposure to PM_2.5_ collocated the low-cost sensors with ambient regulatory monitoring stations or some gold-standard instruments for a long period to obtain the calibration model before the personal monitoring began ^21, 24, 30–33, 37, 38, 41^. This becomes impractical if there is a large number of low-cost sensors, and challenging when these sensors are used in a different environment with different particle composition. Recently, some in-situ calibration approaches have been proposed to address this problem ^52–56^. Other personal monitoring studies collocated low-cost sensors with gravimetric filter-based sampling on site, and obtained a correction factor by comparing the integrated PM_2.5_ mass concentration from the sensor with that from the filter sample across the entire monitoring period ^20, 36, 42, 44, 45, 48, 57^. Nevertheless, this approach loses high-resolution real-time information with large variation from the sensors, and only uses the long-term average.

To address these challenges, this study employed low-cost sensors and GPS receivers integrated into a single wearable device to conduct a two-week personal monitoring of PM_2.5_ exposure for 168 adults recruited from the Washington State Twin Registry between 2018 and 2021. The goals of this study were to (1) quantify the in-situ calibrated personal exposure of PM_2.5_ of participants, partitioned across microenvironments identified using an automated spatial merging method, and (2) compare and contrast this personal monitoring approach with the more common approach of assessing exposure based on ambient concentration at the residential location.

## 2 Methods

### 2.1 Study design

Participants were monozygotic (MZ) twins living in Washington, USA, who were recruited from the Washington State Twin Registry ^58, 59^ for a study that investigated associations between personal exposure monitoring (including PM_2.5_ and allergens) and health ^60, 61^. The present study only focused on the personal exposure monitoring aspects of the parent study. Exclusion criteria included residence outside of Washington state, living with a co-twin, physical limitations that limited mobility, pregnancy, smoking or regular secondhand exposure to tobacco smoke, and regular use of NSAID medications. A total of 168 adult twins were finally recruited between April 2018 and June 2021. Each participant carried the personal monitoring device for two weeks, as well as a stand-alone GPS monitor (details in Section 2.2). The local Institutional Review Board approved this study, and all participants provided informed consent (WSU IRB #18773). Details for study design are available in SM1 **Section S1**.

### 2.2 Data collection

We developed a Portable University of Washington Particle (PUWP) monitor to obtain the personal time and location-specific exposure to PM_2.5_. The PUWP utilizes a low-cost optical light-scattering real-time particle sensor (Plantower PMS A003), which provides multiple channels of particle measurement information: counts of particles for six size bins (i.e., >0.3 μm, >0.5 μm, >1 μm, >2.5 μm, >5 μm, and >10 μm), which were logged at 1-minute intervals. The number concentrations from this sensor have been validated to have a good linear relationship with the reference instrument, and have been applied to indoor, outdoor, and personal monitoring ^38, 62–64^. The Plantower sensor additionally provides the mass concentration of particles with three size bins (i.e., <1 μm, <2.5 μm, and <10 μm), based on the particle number concentrations (PNC) and the sensor manufacturer’s proprietary algorithm. The PUWP was also equipped with sensors for temperature and relative humidity (Honeywell HIH6131-021-001) and a GPS receiver (Adafruit 790 Ultimate GPS Module with MTK3339 Chipset) to record the real-time coordinates at the same frequency as PM_2.5_ monitoring. All monitoring data were time-stamped in UTC and stored in a removable microSD memory card. The model of the PUWP is shown in Supplementary Material (SM) 1 **Figure S1**. Each participant was asked to continuously wear the PUWP monitor for two weeks. Each pair of twins was required to perform the personal PM_2.5_ monitoring over roughly the same time period (e.g., starting within one week of each other).

### 2.3 Data processing

The data processing procedure is shown in **Figure 1**, which was divided into four steps, including data pre-processing, context identification, calibration of air pollution data, and exposure assessment.

**Figure 1.**
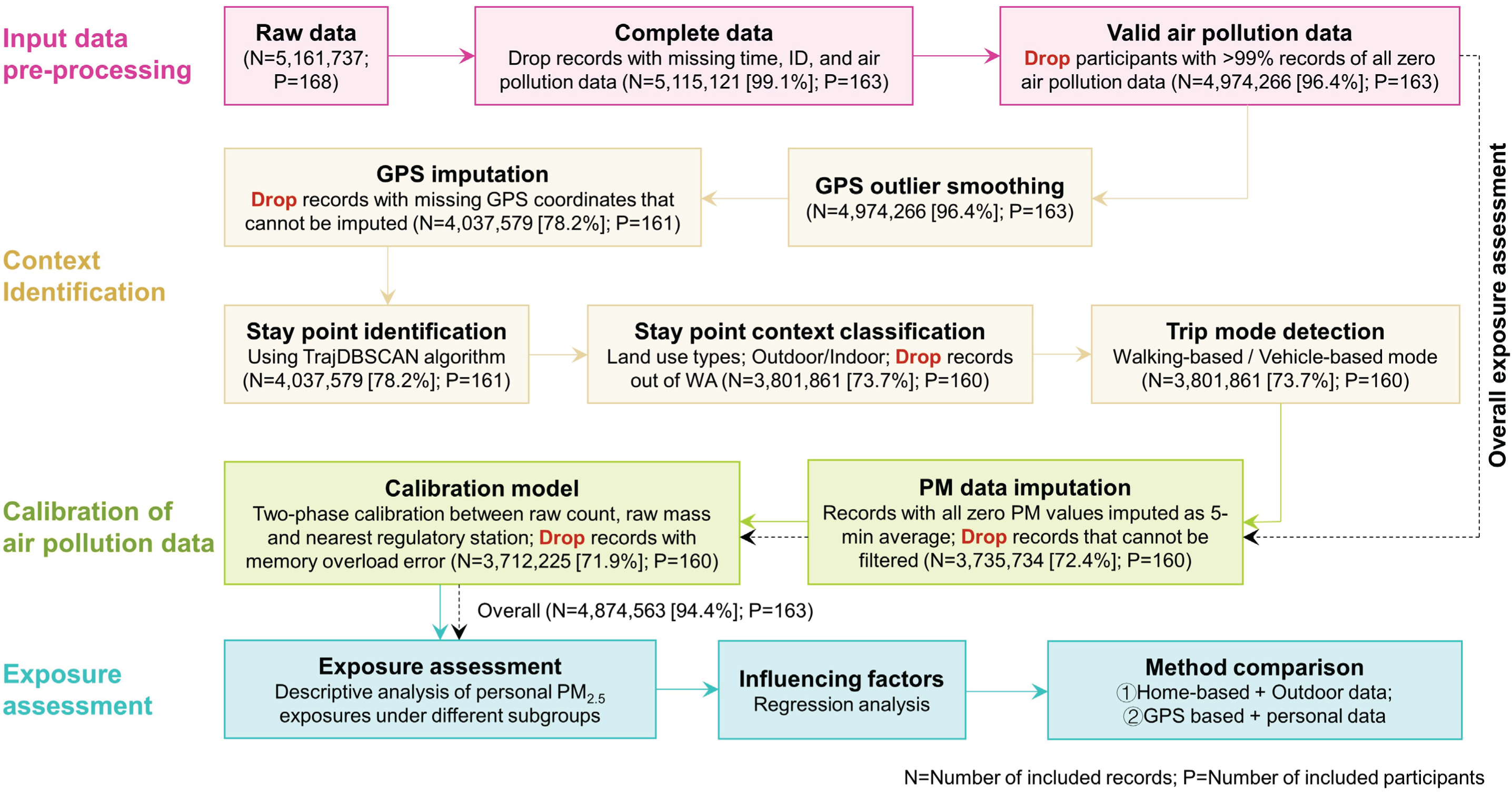
Flowchart of the data processing of personal exposure monitoring.

#### 2.3.1 Data pre-processing

The raw dataset has 5,161,737 data points in total, collected from 168 participants. We first dropped data points with missing timestamps, ID, or air pollution data (*N* = 46,616, 0.9%). We then dropped the participants if their PNC and mass concentration were zero values for over 99% of the monitoring time (*N* = 140,855, 2.7%). For the overall exposure assessment, the remaining data points (*N* = 4,974,266, 96.4%) from 163 participants were directly input into the calibration step (Section 2.3.3), rather than through the context identification step (Section 2.3.2).

#### 2.3.2 Context identification

The collected GPS coordinate data were first processed through a moving median filter and missing data imputation ^65^. After excluding GPS data that cannot be imputed, the remaining data points with GPS (*N* = 4,037,579, 78.2%) were then used to identify the corresponding microenvironments. We first applied the TrajDBSCAN clustering algorithm to classify all data points into different clusters of stay points (i.e., locations where an individual stays for a period of time) and trips ^66–69^. Based on land use data from the Washington State Geospatial Portal ^70^, the land use type of the nearest land use polygon was assigned to each cluster of stay points. We used a 10-m buffer of the building polygons from OpenStreetMap to identify whether the stay points were located in indoor environments. Since study recruitment occurred within Washington state, we excluded those stay points which were out of Washington State (*N* = 235,718, 4.6%). For data points detected as trips, we followed the method of Yi et al., and used the mean and standard deviation of speed and travel distance for each trip to classify all trips into vehicle-based trips and walking-based trips ^65^. All the vehicle-based trips were assigned as the vehicle microenvironment and considered as indoor activities, while the walking-based trips were assumed to be outdoor and assigned as the land use type of the nearest land use polygon. Details for context identification are available in SM1 **Section S2**. There was a total of 3,801,861 data points (73.7%) from 160 participants left after context identification.

#### 2.3.3 Calibration of air pollution data

We used an in-situ calibration approach to compare the hourly average sensor data when the microenvironment context identifies that the sensor is outdoors with the monitoring data from regulatory monitoring stations. We assumed that the outdoor PM_2.5_ mass concentration could be estimated using the nearest regulatory monitoring data if the nearest regulatory monitoring station was relatively close to the outdoor sensor location. In this study, we tried different cut-off distances ranging from 0.5 to 5 km, to maximize the goodness-of-fit of the linear mixed model for this calibration. Details for calibration methods are available in SM1 **Section S3**.

#### 2.3.4 Exposure assessment

Two exposure metrics were used in this section, including exposure concentration and exposure dose. The exposure concentration is the PM_2.5_ mass concentration at a given time (in μg/m^3^), while the exposure dose refers to total mass of PM_2.5_ inhaled during the two-week monitoring period (in μg).

Although data for this study were collected from twin pairs, we assessed individual-level mean hourly PM_2.5_ exposure concentrations during the two-week monitoring period. This part of the analysis did not rely on the microenvironment identification, so we used as many eligible data points as possible (i.e., methods described in Sections 2.3.1 and 2.3.3). A total of 4,874,563 data points (94.4%) from 163 participants were included. Personal exposure concentrations of PM_2.5_ were compared among different demographic and socioeconomic status (SES) characteristics, including age, sex, race, marital status, highest education level, and annual household income.

Next, we assessed the spatiotemporal patterns of the personal exposures based on microenvironment context identification (methods in Section 2.3.2), with a total of 3,712,225 data points (71.9%) from 160 participants included. In order to obtain the more accurate time spent in different microenvironments and contribution of each microenvironment to the total exposure, we excluded the invalid participant-days which had less than 6 hours for one participant ^65^. From the spatial perspective, personal exposure concentrations of PM_2.5_ in eight microenvironments (including seven land use types and vehicles) and indoor/outdoor environments were summarized across the valid days in the two-week monitoring period and compared with each other. To evaluate the contribution of each microenvironment to the cumulative exposure, the proportion of exposure dose for the *k*th microenvironment was further calculated. From the temporal perspective, we compared the personal PM_2.5_ exposure concentrations in different seasons and hours in a day.

In addition, for comparisons with the personal exposure monitoring data, we assessed the hourly average exposure concentrations at participants’ residential location based only on regulatory monitoring station data, which is an approach used in many epidemiological studies. We applied the inverse distance weighted (IDW) interpolation to obtain the outdoor hourly average PM_2.5_ concentrations at participants’ home addresses from the regulatory monitoring network data ^71–75^. Details for exposure assessment are available in SM1 **Section S4**.

### 2.4 Statistical analysis

For the descriptive analysis, boxplots and a series of summary statistics, including mean, standard deviation, 2.5^th^, 25^th^, 50^th^, 75^th^, and 97.5^th^ percentiles, minimum, and maximum, were used to summarize the distribution of personal exposure concentrations overall or in different subgroups across the two-week monitoring period. As the exposure distributions were long-tailed and non-normal, the Wilcoxon test and Kruskal-Wallis test were used to compare the exposure concentrations between two subgroups and across multiple (≥3) subgroups, respectively. If there was a significant difference after the Kruskal-Wallis test, a Bonferroni adjustment was used for the post-hoc pairwise comparison. Furthermore, a linear mixed model was applied to estimate the associations of multiple spatiotemporal factors with personal exposure to PM_2.5_.

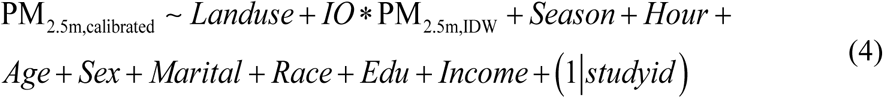

where *Landuse* represents seven dummy variables for land use types (reference: other land use); *IO* is the dummy variable for indoor or outdoor environments (reference: outdoor); PM_2.5m,IDW_ is the estimated outdoor PM_2.5_ concentrations at participants’ home addresses through IDW interpolation, which represents the regional background concentration for this participant, μg/m^3^; *Season* represents three dummy variables for seasons (reference: spring); *Hour* represents 23 dummy variables for hours in a day (reference: 0 o’clock); *Age* represents four dummy variables for age groups (reference: 0-29 years old); *Sex* is the dummy variable for sex (reference: female); *Marital* is the dummy variable for marital status (reference: unmarried); *Race* is the dummy variable for race (reference: non-white); *Edu* represents two dummy variables for highest education level (reference: lower than bachelor); and *Income* is the dummy variable for annual household income (reference: low income). To better interpret the possible impacts of all categorical variables in the above model, the estimated marginal means (EMMs) were used for each level of each categorical variable ^76^. The 95% confidence intervals of coefficients in the linear mixed model were obtained through the profile likelihood approach ^77^. All data analysis in the “Methods” section were performed in R V4.2.2 software using lme4, lmerTest, sf, sp, geosphere, lubridate, reshape2, and emmeans packages ^78–87^, as well as QGIS V3.16.7.

## 3 Results

### 3.1 Overall results of personal PM_2.5_ exposure levels

The calibration results show a high correlation between PM_2.5_ hourly average concentration predictions and those from the nearest regulatory monitoring stations (R^2^ = 0.93, RMSE = 0.1 μg/m^3^), suggesting the high performance of the in-situ calibration in this study (details in SM1 **Section S3**). After the calibration, the average personal PM_2.5_ exposure concentration of all data points was 8.1±15.8 μg/m^3^, and the median exposure concentration reached 4.6 μg/m^3^. **Figure 2** shows the boxplots of individual-level personal PM_2.5_ exposure concentration s grouped by twin pairs. A total of 163 participants and 78 complete twin pairs were included. Most participants were exposed to PM_2.5_ concentrations lower than 10 μg/m^3^ with relatively small variation, while some participants had a very high PM_2.5_ exposure with substantial variation, such as AIR1069B, AIR1796B, and AIR4602B. Detailed summary statistics of personal exposure concentrations for all participants are provided in **Table S5**.

**Figure 2.**
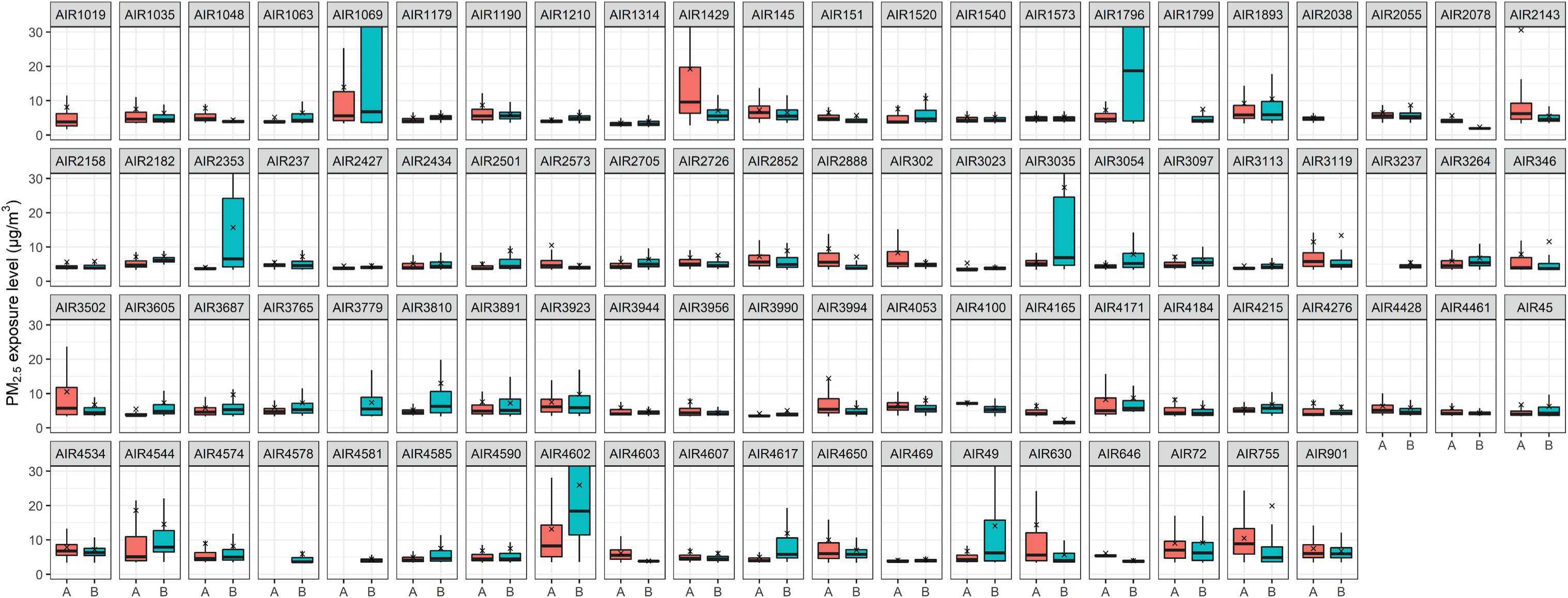
Boxplots of personal PM_2.5_ exposure concentration for all participants grouped by twin pairs. **Note:** Each box shows the 1^st^/3^rd^ quartile (Q1/Q3) at the lower/upper end of the box, and the median as a horizontal line inside the box. The whiskers extend to the smallest and the largest values within 1.5 × interquartile range (IQR) from Q1 and Q3. The red cross represents the mean value. A (red) and B (green) represent two individuals in a pair of twins. There are some missing boxplots because the personal monitoring data of these participants were dropped during the data processing. Finally, there are 163 participants and 78 complete twin pairs.

The personal PM_2.5_ exposure concentrations were also compared for various demographic and SES characteristics. **Figure S8 (*a*)** shows the boxplots of median participant-level exposures. Most medians were lower than 7.5 μg/m^3^. Significant differences were only found between the 50-59 and over 60 years age groups, and between currently married and unmarried participants. The exposure concentrations of participants over 60 years old were lower than the 50-59 age group (difference = 0.9 μg/m^3^). The exposure concentrations of currently married participants were also lower than those of unmarried participants (difference = 0.4 μg/m^3^). **Figure S8 (*b*)** provides the boxplots of 97.5^th^ percentiles of participant-level exposures. We found that the 97.5^th^percentile of exposure concentrations for females were significantly higher than that for males (difference = 5.4 μg/m^3^), while no significant differences were detected for other covariates. Detailed summary statistics can be found in **Table S6**.

### 3.2 Spatiotemporal patterns of personal PM_2.5_ exposures

In this section, we investigated the possible effect of spatiotemporal factors on personal PM_2.5_ exposure variability. Let’s first consider one participant, AIR4585A, as an example to observe the influence of spatiotemporal factors on PM_2.5_ exposure concentrations. **Figure S9** illustrates the 1-minute PM_2.5_ exposure concentration of AIR4585A along with the GPS position for two days in April, 2018. On the first day, this participant stayed at home until about 6 am. Then, they took a vehicle-based trip to a shopping center and finally drove back home around 5:30 pm. The personal exposure concentrations did not vary a lot during this day and ranged from 3.4 to 7.8 μg/m^3^. The highest exposure concentrations occurred in the shopping center and residence. On another day, they stayed at home for most of the day, and only drove to a location in the east, and back home between 4 and 5 pm. The personal exposure concentration ranged from 3.5 to 26.7 μg/m^3^, and the highest exposure was observed at around 6 am in the residential microenvironment. This example reveals that the large variation in this individual’s personal PM_2.5_ exposures was related to spatiotemporal factors, such as time spent in various microenvironments and time of day.

From the perspective of space, different microenvironments can play a critical role in the spatiotemporal variation in personal PM_2.5_ exposure levels. **Figure S10** shows the time pattern in different microenvironments. Participants spent 78% of their time in indoor environments, consistent with previous survey results ^9, 10^. Participants also spent most of their time in residential land use, accounting for 67% of total time, followed by public facilities (9%) and commercial land use (7%). **Figure 3** and **Table S7** show the personal exposure concentrations in different microenvironments using the original 3.7 million data points. Differences between all subgroups were found to be significant (*p* < 0.001). The highest median exposure concentration occurred in industrial land use (5.7 μg/m^3^), while the lowest was in public facilities land use (4.3 μg/m^3^). Median indoor exposure concentration was comparable to the median for outdoor exposure (4.6 μg/m^3^). However, all mean values were higher than the 75^th^ percentile, suggesting that these exposure concentrations followed a right-skewed distribution pattern with some extremely high concentrations. For example, although the median exposure concentration in the residential land use was 4.6 μg/m^3^, the mean was 8.3 μg/m^3^ and the proportion of exposure concentrations higher than 20 μg/m^3^ (95^th^ percentile of all data points) reached 5.2%. Combining the time pattern and exposure concentrations, the contribution of exposure dose in different microenvironments can be further estimated, shown in **Figure 4**. Since the exposure concentrations had no substantial difference among various microenvironments, the contribution of exposure dose is nearly proportional to the time spent in each kind of microenvironment. Specifically, indoor exposure dose contributed 77% to total exposure dose, which was much higher than outdoor exposures. Additionally, the contribution of residential exposure dose ranked first among all land use types and accounted for 69% of total exposure dose.

**Figure 3.**
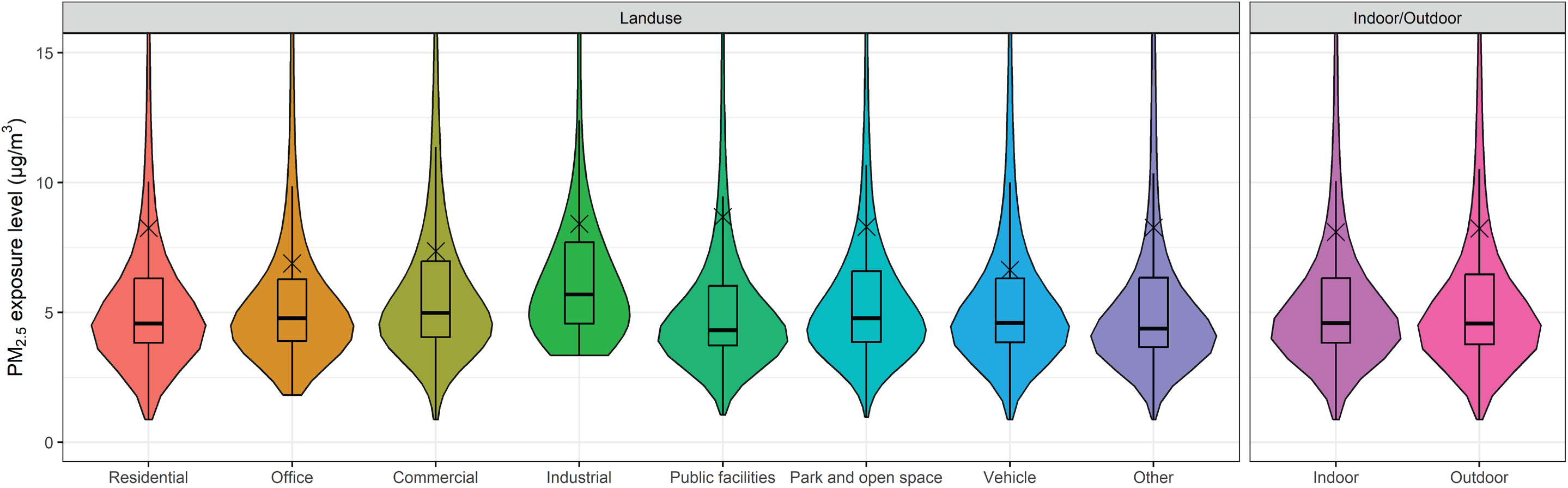
Violin plots of personal PM_2.5_ exposure concentrations in different microenvironments.

**Figure 4.**
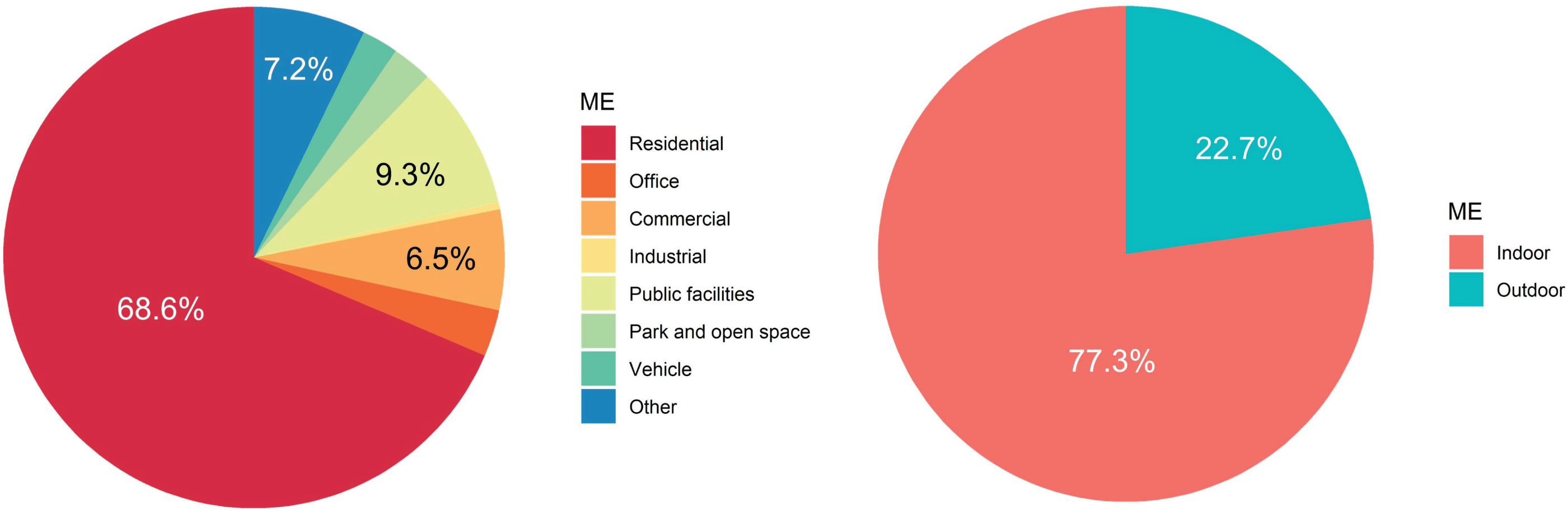
Proportion of total PM_2.5_ exposure dose in different microenvironments.

From the perspective of time, personal exposures were compared under two temporal scales, i.e., hourly diurnal patterns and variations across participants monitored in different seasons, as shown in **Figure 5** and **Table S8**. The personal PM_2.5_ exposure concentration in winter was 12.2±26.3 μg/m^3^, which was higher than other seasons. Diurnal exposure patterns illustrate that for this study, participants’ personal PM_2.5_ exposure tended to rise at 6 pm and reach a peak at 7 pm (10.9±21.5 μg/m^3^), then gradually decrease until 5 am the next day (6.6±10.8 μg/m^3^, the lowest level).

**Figure 5.**
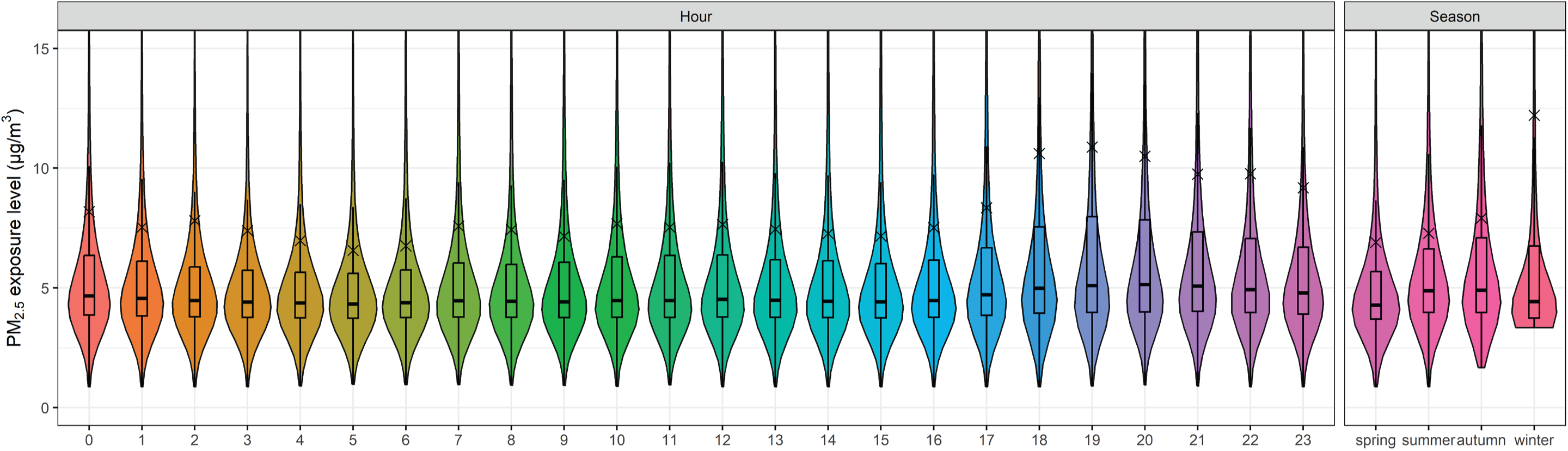
Violin plots of personal PM_2.5_ exposure concentrations in different seasons and hours in a day.

### 3.3 Comparison with home-based epidemiologic approach

The exposure concentration based on the residential location assessment approach with IDW interpolation of the regulatory monitoring station data was 6.1±7.2 μg/m^3^, which was lower than the GPS-based personal monitoring results (8.1±15.8 μg/m^3^). The variation in exposure concentrations from the residential location approach was also smaller than that in personal exposure concentrations measured in this study. We provide two examples representing two different scenarios to support the above statement. As is shown in **Figure S11 (*a*)**, the personal exposure concentration had a more dramatic fluctuation in the GPS-based approach, which was influenced by many indoor exposure peaks, than the residential location assessment method. Therefore, the residential location exposure assessment may underestimate the personal exposure method for PM_2.5_. The Pearson correlation between the two results was only 0.28. In contrast, in **Figure S11 (*b*)**, in August, 2018, the residential location assessment approach overestimated the personal exposure concentration of the participant. The high outdoor PM_2.5_ concentrations were likely caused by wildfires that occurred during the summer, and reached over 50 μg/m^3^ on some days. However, the personal exposure concentration was lower than 25 μg/m^3^ for most of the time, except for some peaks, leading to a moderate Pearson correlation (r = 0.48). The above evidence demonstrates that high outdoor PM_2.5_ concentrations at home addresses does not necessarily mean high personal exposure concentrations, and vice versa.

### 3.4 Regression analysis of multiple spatiotemporal factors

The subgroup analysis in Section 3.2 and 3.3 considers different influencing factors separately, and does not account for potential confounding. However, spatiotemporal factors along with demographic and SES covariates can affect personal exposure concentrations simultaneously. **Table 1** shows the coefficients of the linear mixed model with multiple independent variables. From the perspective of microenvironments, the positive main effect of *IO* suggests that indoor exposure concentration was higher than outdoor exposure by 0.5 μg/m^3^ on average. In addition, the highest PM_2.5_ exposures occurred in the industrial and residential land uses, higher than the lowest land use (office) by 3.2 and 3.0 μg/m^3^, respectively. The personal exposure in park and open space microenvironments, as well as the vehicle microenvironment, was also relatively low.

**Table 1.** Results of the linear mixed model with multiple spatiotemporal covariates.

| Variable | Estimated Marginal<br>Mean ($\mu\text{g}/\text{m}^3$) | Coefficient | 95% Confidence Interval |
| --- | --- | --- | --- |
| **(Intercept)** | — | 8.545 | (5.581, 11.510) * |
| **IO** |  |  |  |
| Outdoor | 5.800 | Ref | Ref |
| Indoor | 5.297 | 0.520 | (0.464, 0.575) * |
| **PM<sub>2.5m,IDW</sub>** | — | 0.279 | (0.275, 0.284) * |
| **IO × PM<sub>2.5m,IDW</sub>** | — | -0.168 | (-0.173, -0.163) * |
| **Land use type** |  |  |  |
| Other land use | 6.154 | Ref | Ref |
| Commercial | 5.830 | -0.324 | (-0.418, -0.230) * |
| Industrial | 6.726 | 0.571 | (0.329, 0.814) * |
| Office | 3.511 | -2.643 | (-2.758, -2.528) * |
| Park and open space | 5.391 | -0.763 | (-0.882, -0.645) * |
| Public facilities | 5.808 | -0.346 | (-0.438, -0.254) * |
| Residential | 6.511 | 0.357 | (0.278, 0.435) * |
| Vehicle | 4.461 | -1.694 | (-1.805, -1.582) * |
| **Season** |  |  |  |
| Spring | 6.740 | Ref | Ref |
| Summer | 5.429 | -3.905 | (-4.004, -3.806) * |
| Autumn | 2.835 | -1.312 | (-1.513, -1.111) * |
| Winter | 7.191 | 0.451 | (0.308, 0.594) * |
| **Hour in a day** |  |  |  |
| Hour 0 | 5.712 | Ref | Ref |
| Hour 1 | 5.014 | -0.698 | (-0.802, -0.594) * |
| Hour 2 | 5.165 | -0.547 | (-0.651, -0.443) * |
| Hour 3 | 4.677 | -1.035 | (-1.140, -0.931) * |
| Hour 4 | 4.264 | -1.448 | (-1.552, -1.344) * |
| Hour 5 | 3.853 | -1.859 | (-1.963, -1.755) * |
| Hour 6 | 4.042 | -1.670 | (-1.774, -1.566) * |
| Hour 7 | 4.881 | -0.831 | (-0.935, -0.727) * |
| Hour 8 | 4.735 | -0.977 | (-1.082, -0.873) * |
| Hour 9 | 4.485 | -1.227 | (-1.331, -1.123) * |
| Hour 10 | 5.018 | -0.694 | (-0.798, -0.590) * |
| Hour 11 | 4.889 | -0.823 | (-0.927, -0.719) * |
| Hour 12 | 5.100 | -0.613 | (-0.716, -0.509) * |
| Hour 13 | 5.001 | -0.711 | (-0.815, -0.608) * |
| Hour 14 | 4.871 | -0.841 | (-0.945, -0.738) * |
| Hour 15 | 4.855 | -0.858 | (-0.961, -0.754) * |
| Hour 16 | 5.195 | -0.517 | (-0.620, -0.414) * |
| Hour 17 | 5.997 | 0.285 | (0.182, 0.387) * |
| Hour 18 | 8.097 | 2.385 | (2.282, 2.487) * |
| Hour 19 | 8.327 | 2.614 | (2.512, 2.717) * |
| Hour 20 | 7.925 | 2.212 | (2.110, 2.315) * |
| Hour 21 | 7.190 | 1.478 | (1.375, 1.581) * |
| Hour 22 | 7.190 | 1.477 | (1.374, 1.581) * |
| Hour 23 | 6.688 | 0.975 | (0.872, 1.079) * |
| **Age group** |  |  |  |
| 0-29 [20] | 9.175 | Ref | Ref |
| 30-39 [77] | 5.058 | -4.116 | (-7.481, -0.751) * |
| 40-49 [13] | 3.962 | -5.213 | (-9.750, -0.675) * |
| 50-59 [32] | 6.460 | -2.714 | (-6.457, 1.028) |
| $\geq 60$ [9] | 3.089 | -6.085 | (-11.780, -0.391) * |
| **Sex** |  |  |  |
| Female [120] | 6.491 | Ref | Ref |
| Male [31] | 4.607 | -1.884 | (-4.686, 0.918) |
| **Marital status** |  |  |  |
| Unmarried [62] | 5.503 | Ref | Ref |
| Married [89] | 5.594 | 0.091 | (0.026, 0.156) * |
| **Race** |  |  |  |
| Non-white [9] | 4.011 | Ref | Ref |
| White [142] | 7.087 | 3.076 | (2.884, 3.268) * |
| **Highest education level** |  |  |  |
| Lower than BA [31] | 5.065 | Ref | Ref |
| BA [66] | 6.747 | 1.682 | (1.554, 1.809) * |
| Higher than BA [54] | 4.834 | -0.231 | (-0.375, -0.088) * |
| **Annual household income** |  |  |  |
| Low [54] | 5.535 | Ref | Ref |
| High [97] | 5.563 | 0.028 | (-0.036, 0.092) |

Notes: \* refers to $p < 0.05$ . Ref means the reference level for the categorical variable. The number in brackets for age group, sex, marital status, race, highest education level, and annual household income refers to the number of participants included in each category.

From the perspective of time, highest exposure concentration was found in winter, which is consistent with findings in Section 3.4, while the lowest exposure was in summer. The exposures between 7 pm and 8 pm contributed the highest to total personal PM_2.5_ exposure in a day, while 5 am contributed the lowest. This result agrees well with previous subgroup analysis. Generally, the personal exposure concentration gradually decreased from 7 pm to 5 am and from 8 am to 3 pm.

We included the outdoor concentration at home addresses (PM_2.5m,IDW_) in this regression model to represent the regional background levels. The positive coefficient of 0.279 suggests that the personal exposure was weakly but positively correlated with the outdoor concentrations. The significant negative interaction between indoor/outdoor environment and regional background PM_2.5_ (–0.168) reveals that the correlation between indoor personal exposure and outdoor concentrations at the home addresses (i.e., 0.279-0.168=0.111) was much lower than that between the outdoor personal exposure and outdoor home-based concentrations.

Some demographic and SES covariates were also significant in this regression analysis after adjustment by the above spatiotemporal factors. Participants younger than 30 years old had the highest personal exposure, while those older than 60 years old had the lowest. Exposure concentrations among males were not significantly different from female levels. Unmarried, non-white participants with an education level higher than BA had significantly lower exposure concentrations to PM_2.5_.

## 4 Discussion

This study assessed the personal PM_2.5_ exposure levels of 163 participants over two weeks of monitoring, using automatically identified microenvironments and an in-situ calibration approach. The calibration approach showed good performance against the criterion measure (R^2^ = 0.93, RMSE = 0.1 μg/m^3^). Mean (SD) and median (IQR) personal PM_2.5_ exposure concentrations were 8.1 (15.8) μg/m^3^ and 4.6 (2.5) μg/m^3^. Overall, we found that exposure levels varied by certain spatiotemporal characteristics. Notably, based on multivariate regression analysis, indoor exposure concentrations were higher than outdoor exposures, after controlling for other confounding factors. Exposure concentrations that occurred in industrial and residential land uses were higher than in other land use categories. Additionally, personal exposures were the highest during winter and evening hours (around 7 pm). Finally, the residential location exposure assessment method based on interpolation of regulatory monitoring station measurements that is used in many epidemiological studies did not capture the spatiotemporal variations in personal PM_2.5_ exposures observed for participants in this study.

Different microenvironments contributed to the large variation in spatiotemporal personal exposure concentration to PM_2.5_. This study found that exposure concentrations occurring in industrial and residential land use were the highest among all microenvironments. A few participants (e.g., AIR1796B, AIR2353B, and AIR3035B) experienced extremely high PM_2.5_ exposure concentrations (>100 μg/m^3^) in their residences during 7 am, 10-11 am, and 8-10 pm (AIR1796B), 5-6 pm (AIR2353B), and 4-6 pm (AIR3035B), respectively. Several previous studies also observed a relatively high exposure concentration in residences compared to other microenvironments ^20, 21, 34^. Liu et al. demonstrated in their personal monitoring study of pregnant women that there were more peaks and higher peak exposure concentrations in home residential locations ^57^. This is likely due to indoor cooking and heating activities, which is supported by the finding that higher exposures were observed around 7 pm (i.e., dinner time, also found by Koehler et al. ^20^) and winter (i.e., residential heating season). Cooking oil fumes can lead to substantial particle exposures, even with clean fuel (such as electricity), so cooking is an important indoor source of PM_2.5_ which cannot be ignored ^12, 88, 89^. Another piece of indirect evidence on the contribution of cooking is that several studies reported relatively high PM_2.5_ exposures in restaurants or eatery microenvironments, which were also likely affected by cooking ^30, 42, 45^. For heating, estimates from the 2021 American Community Survey (ACS) show that there are a considerable number of households in Washington State which use wood in their residences ^90^. Some field studies have found that solid fuel users are exposed to a significantly higher PM_2.5_ concentration in their residences than clean fuel users ^32, 33, 91^. Together, the above evidence can explain the high exposure concentration in residential land uses, and at 7 pm and in winter. We also notice that 70% of exposure at 7 pm with the highest exposure concentration occurred indoors, which further emphasizes the substantial contribution of indoor exposure to the total personal exposure.

In addition to exposure concentration, exposure dose in residential land use is also of great importance. Lin et al. estimated the contribution of microenvironments to the total exposure dose and found that residential exposures accounted for 74.7% of total exposures across all seasons ^45^, which is very close to the 68.6% estimate found in this study. Li et al. found that for retired adults in two megacities in China, residential microenvironments accounted for about 85% of the total PM_2.5_ exposure dose ^42^. The slightly higher contribution is likely due to different time-activity patterns between retired people and adults across different age groups in this study. Liu et al. revealed that the peak exposure dose (i.e., area under the curve (AUC) at peaks) in residences was higher than other microenvironments ^57^. Therefore, from the perspective of both exposure level and exposure dose, PM_2.5_ exposure in residential microenvironments should be emphasized, and controlling these exposures should be prioritized in the future.

Ventilation and air purification are commonly used strategies to control the residential PM_2.5_ exposure. Previous studies show that using portable air cleaners (PACs) with high-efficiency particulate air (HEPA) filters can substantially reduce the exposure level of PM_2.5_ in households even with uncontrolled ventilation conditions ^92–95^. Liu et al. focused on different intervention strategies to mitigate cooking-related PM_2.5_ exposure, and found that combining PACs and ventilation was the most effective way in removing cooking-related PM_2.5_ ^96^. They also suggested that adding a stove hood was very helpful in reducing PM_2.5_ concentrations due to cooking ^96^. Some studies further considered both health benefits due to exposure reduction and control costs to provide the optimal control approach, such as the best ventilation rate and concentration threshold for PACs ^97–99^. Besides, source control is another efficient strategy for reducing residential PM_2.5_ levels, which includes avoiding smoking indoors, reducing solid fuel use for cooking and heating, and replacing old stoves with high-efficiency ones ^100^. Some studies pointed out the effect of cooking method and oil types on PM_2.5_ emission rates, where pan-frying and stir-frying emit more particles than deep frying, steaming, and boiling, and olive oil generates more particles than other oil types such as peanut and sunflower oil ^101–103^. These findings suggest that change of cooking habits if possible can also reduce residential PM_2.5_ exposure concentrations.

This study’s findings related to the importance of PM_2.5_ exposures in industrial land use settings have not been well-documented in previous studies. Although some previous personal monitoring studies have investigated personal exposure concentrations in workplaces generally ^20, 21, 30, 45^, but not specifically for industrial land use. Over 60% of exposure data in the industrial land use in this study happened outdoors, so the high exposure concentrations in industrial areas may be more related to outdoor PM_2.5_ pollution. The outdoor PM_2.5_ concentrations in industrial areas can be contributed from both industrial emissions and freight transport emissions, such as gasoline/diesel vehicles, trains, and ships, which can be supported by inventories and source apportionment studies. The Washington comprehensive emission inventory in 2020 provided the source contributions to PM_2.5_ in King County, which was the most urbanized area in Washington and where most participants were located in this study^104^. Industrial/commercial/institutional fuel use, paved and unpaved road dust, on-road mobile sources, point sources, ships, and railroads contributed 8.3%, 9.1%, 4.4%, 0.8%, 0.6%, and 0.2%, respectively. Additionally, a source apportionment study in Beacon Hill (near the industrial district) in Seattle, identified that gasoline/diesel mobile sources and industry contributed 44% and 7% to local PM_2.5_ concentrations ^105^. Another study reported that freight transport (gasoline, diesel, and fuel oil) and industry (metal processing and cement kiln) accounted for 22% and 11% at Duwamish site in the industrial district of Seattle ^106^. It was also found at Duwamish that secondary nitrate, likely related to vehicle emissions, and secondary sulfate, likely associated with industrial emissions, contributed 24% and 20% of PM_2.5_ mass concentrations ^106^. All above evidence suggest that freight transport and industrial emission contribute to the higher personal PM_2.5_ exposure concentration in industrial land use in this study.

The spatiotemporal factors likely explain the variation in personal exposure to PM_2.5_ within participant, which was so large that it exceeded the between-participant variation. A variance component analysis was performed using a random intercept model (details in **Section S5**) ^20^. The intraclass correlation coefficient (ICC) was only 0.20, suggesting a relatively small between-participant difference, compared to the within-participant variation. On one hand, although the median difference between each twin pair was only 0.5 (95% UI: 0.0-5.0) μg/m^3^, health impacts of PM_2.5_ are sensitive to these small changes in exposures at the low exposure range. For instance, according to exposure-response relationships from the GBD Study, if PM_2.5_ concentration increases from 5 to 10 μg/m^3^, the relative risk of ischemic heart disease for people 40-44 years old can increase from 1.20 to 1.36, which means a greater than 13% higher risk of developing these cardiovascular diseases ^107^. Therefore, it is still worthwhile to investigate the impact of this small exposure difference on health outcomes in future epidemiological studies. On the other hand, the large within-participant exposure variation implies that the median or mean exposure concentrations cannot fully represent the whole exposure distribution. Future epidemiological studies can consider other summary statistics, such as 75^th^ and 97.5^th^ percentiles, to depict the peak exposures.

A discrepancy between exposure estimates from the personal monitoring approach and the residential location exposure assessment method based on interpolation of regulatory monitoring station measurements was observed in this study. The residential location exposure assessment approach can sometimes miss some exposure peaks. This may include emissions from some microenvironment-specific sources, such as cooking and smoking, which cannot be reflected by the IDW interpolation of outdoor monitoring data. On the other hand, for outdoor wildfire scenarios, the residential location exposure assessment approach was found to overestimate personal exposures. This can possibly be explained by the participant staying in indoor environments throughout the day, while the outdoor PM_2.5_ had a relatively low infiltration factor indoors, or this participant utilized air cleaners. The isolated peaks seen in the personal monitoring method may result from personal behaviors like window-opening behaviors or going outdoors, or some indoor source emissions. Therefore, the residential location exposure assessment approach used in many epidemiological studies in some cases, may not capture spatiotemporal variations in personal exposure and could underestimate the personal exposure concentration (6.1 vs 8.1 μg/m^3^), which may be the source of bias in air pollution epidemiologic studies. Some previous studies also investigated the comparison between personal exposure and outdoor PM_2.5_ concentration at residential locations ^20, 24–26^. As observed in our study, they also found that the residential location exposure assessment method does not accurately reflect the true exposure concentration. In this study, the median ratio of personal exposure to outdoor residential location-based exposure (denoted as P/O ratio) was 1.08 (IQR: 0.73-1.61), a bit higher than 0.95 (0.79-1.09) and 0.88 (0.69-1.06), values that were reported in studies from two megacities in China ^42^. Since the outdoor PM_2.5_ concentration was much lower in Washington, US than in China, the contribution of indoor source emissions played a more important role in the indoor exposures and corresponding personal exposures, which may lead to a higher P/O ratio in this study. The Pearson correlation between personal and outdoor residential location-based exposure was only 0.10, lower than 0.30 from Koehler et al. in Colorado, USA ^20^, demonstrating a larger difference between the two exposure assessment results in this study. The possible reason for this lower correlation is that we used minute-level data to estimate the correlation while Koehler et al. used daily average level which smoothed the concentration peaks and fluctuations during each day ^20^. The above exposure errors between personal and residential location-based exposures, which include both Berkson errors and classical errors, can then lead to bias and variance inflation of health effects obtained in epidemiological studies ^108, 109^. Kioumourtzoglou et al. proposed a calibration coefficient for health effect estimate from surrogate exposure (i.e., residential location exposure assessment results) based on paired ambient and personal PM_2.5_ monitoring data in 9 cities ^110^. They found that the calibration coefficient was 0.54 (95% CI: 0.42 – 0.65), suggesting an underestimation of health risks using the residential location-based approach ^110^.

There are several strengths of this study to highlight: First, this study combined GPS data with land use and building data to automatically determine the microenvironments at different times for each participant. This approach can substantially save time and labor compared to time-activity diaries and manually identifying microenvironments. Second, this study used a two-week personal monitoring period, which is longer than previous studies that used one or two days of monitoring, and thus can reflect weekly personal exposures more accurately. Third, this study classified microenvironments into seven different land use types and a ‘vehicle’ environment, while most previous studies focused on fewer microenvironments, such as home and school. We also abandoned the usage of “workplace” because it is so general as to include multiple land use types with quite different exposure characteristics.

However, there are some limitations of this study that should be noted. First, the number of participants and the two-week monitoring period were still limited, suggesting that it should be cautious to generalize the findings to larger population. Second, while GPS tracks can tell us about the real-time position and the corresponding microenvironments of the participants, the specific activities they are engaged in are still unknown. Hence, we can only infer the possible activities or sources which led to the high exposures, such as indoor cooking or heating. To avoid burdensome time-activity diaries, a few current studies have used wearable cameras to record people’s real-time activities for aiding the exposure assessment ^48, 111–113^. Deep learning algorithms can then be applied on the images to identify possible activities in various microenvironments. Source apportionment could also be considered via chemical composition analysis, size-resolved particle monitoring, or multi-pollutant monitoring, to help identify the specific indoor activities ^46^. Third, we used 10-m buffers of buildings to distinguish indoor microenvironments from outdoor microenvironments to minimize the effect of GPS measurement errors. However, it cannot completely rule out the probability of misclassification, especially for indoor cases with poor GPS signals. Future studies can compare and apply GPS receivers with higher accuracy, or combine GPS from smartphones and other devices to double check the GPS coordinates. Fourth, limited data points can be used in the calibration in this study, which only covered the relatively low exposure range and might reduce the generalizability of the calibration equation. Future studies can strengthen this in-situ calibration approach by incorporating collocation data with regulatory monitoring station and other stationary low-cost sensor data such as PurpleAir.

## 5 Conclusions

This study used low-cost sensors to perform two-week personal PM_2.5_ monitoring for 168 adults from the Washington State Twin Registry between 2018 and 2021. We combined GPS information with land use and building data to automate the classification of microenvironments and obtain the spatiotemporal patterns of personal exposure to PM_2.5_. We also developed an in-situ calibration approach for the low-cost sensors and obtained good performance compared to the gold standard (R^2^ = 0.93). The multivariate regression results suggest that indoor exposure concentrations were slightly higher than outdoor exposures, and PM_2.5_ exposure concentrations in residential and industrial land uses were higher than other microenvironments. These two high-exposure scenarios accounted for 77% and 69% of the total exposure dose, respectively, which is consistent with previous studies. In addition, winter and the 6-8 pm time block contributed the most to PM_2.5_ exposure, possibly due to indoor cooking and wood combustion for residential heating. Outdoor wildfire events that occurred during a portion of the monitoring period also caused extremely high personal exposures for some participants. Furthermore, this study demonstrates that the residential location exposure assessment approach used in many epidemiological studies cannot accurately reflect the spatiotemporal patterns of personal exposure concentrations, and will likely lead to bias in estimated exposure concentrations. The findings of this study support the use of low-cost sensors and GPS to improve the precision of personal exposure assessment, and reveal specific microenvironments where PM_2.5_ exposures are high (i.e., residential indoor microenvironments) and can be targeted in future interventions to mitigate the deleterious effects of exposures on health.

## Data Availability

All data produced in the present study are available upon reasonable request to the authors.

## Acknowledgements

We acknowledge that this work was funded by a grant from the National Institute of Health (NIH) NIEHS ES024715. We thank Shelby Tarutis for her work in recruitment and data collection. We thank the twins for their participation in this study.

## Notes

### Competing Interest Statement

The authors have declared no competing interest.

### Funding Statement

This study was funded by a grant from the National Institute of Health (NIH) NIEHS ES024715.

### Author Declarations

Ethics committee/IRB of Washington State University gave ethical approval for this work.

