## Supplementary Material 1 for "Using low-cost sensors and GPS to assess spatiotemporal variations in personal exposure to PM_2.5_ in the Washington State Twin Registry"

[†] Anne V. Moudon passed away during the preparation of this manuscript. We are very grateful to her contribution to GPS-based mobility and its relationship to urban form in this study.

This supplementary material includes 5 sections, 11 figures, and 4 tables.

Table S5-S8 are available in supplementary material 2 in xlsx format.

### Contents

**Section S1 Study design ....................................................................................S3**

**Section S2 Context identification................................................................S5**

S2.1 GPS coordinate processing before context identification............................S5

S2.2 Overview of context identification .............................................................S5

S2.3 Identification of stay points .........................................................................S6

S2.4 Microenvironment (ME) classification of stay points .................................S7

S2.5 Trip mode detection .....................................................................................S12

**Section S3 Calibration of air pollution data .........................................................S13**

S3.1 Methods of calibration .................................................................................S13

S3.2 Results of calibration ...................................................................................S15

**Section S4 Exposure assessment ...........................................................................S20**

S4.1 Methods of exposure assessment ..............................................................S20

S4.2 Results of exposure assessment ................................................................S22

**Section S5 Comparison between within-participant and between-participant variations ..........................................................................................S25**

**References ...........................................................................................................S26**

### Section S1 Study design

Participants were monozygotic (MZ) twins living in Washington, USA, who were recruited from the Washington State Twin Registry [1, 2] for a study that investigated associations between personal exposure monitoring (including $PM_{2.5}$ and allergens) and health [3, 4]. The present study only focused on the personal exposure monitoring aspects of the parent study. Exclusion criteria included residence outside of Washington state, living with a co-twin, physical limitations that limited mobility, pregnancy, smoking or regular secondhand exposure to tobacco smoke, and regular use of NSAID medications. A total of 168 adult twins were recruited between April 2018 and June 2021. Data collection was paused from April 2020 to December 2020 due to the pandemic. Participants who enrolled prior to the pandemic attended an in-person study visit, which included a blood draw, spirometry test, and blood pressure measurement, followed by a detailed explanation of the data collection procedures. Participants who enrolled starting in January 2021 completed study procedures at home. We sent a blood pressure monitor with instructions for obtaining an accurate reading, and materials for collecting blood samples on blood spot cards. Each participant carried the personal monitoring device (i.e., PUWP monitor in Section 2.2) for two weeks, as well as a stand-alone GPS monitor (QStarz BT-Q1000XT) and accelerometer (Actigraph GT3X). They completed three questionnaires: one about their general health and wellbeing, one about their neighborhood, and one about their typical food consumption. The local Institutional Review Board approved this study, and all participants provided informed consent (WSU IRB #18773).

**Figure S1.** Model of the Portable University of Washington Particle (PUWP) monitor. (*a*) Custom motherboard integrates low-cost readily available components: sensors, datalogging, Bluetooth, etc. (*b*) Final appearance of the PUWP monitor.

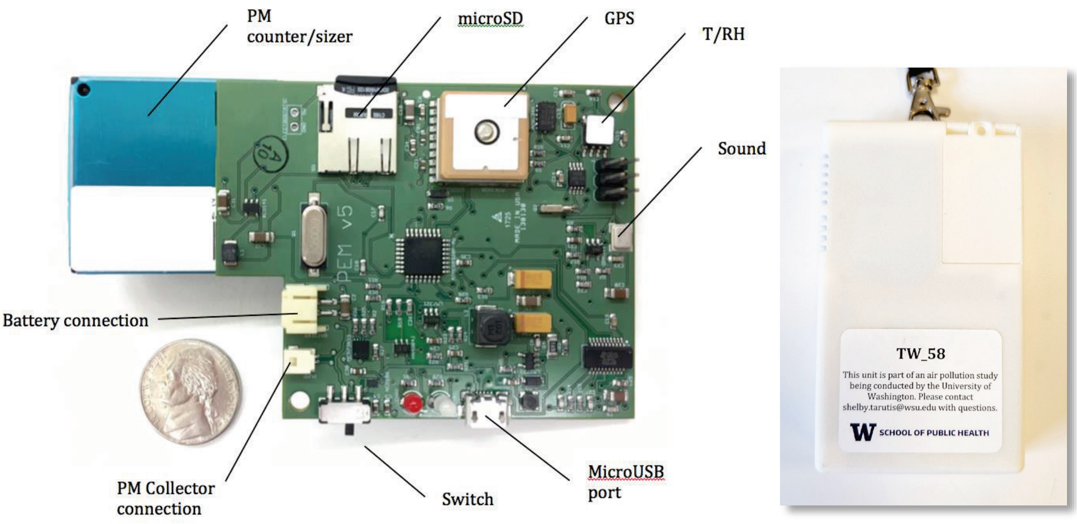


(*a*) (*b*)

### Section S2 Context identification

This section will introduce the detailed methods of context identification in this study.

#### S2.1 GPS coordinate processing before context identification

We first applied a moving median filter to remove GPS outliers in the window of five nearest data points, which is approximately 5 minutes [5]. The outliers were defined as those observations with a distance >2700 m (i.e., 45 m/s or 100 mph multiplied by 60 s) from the median coordinates within the moving window [5]. Then, we attempted to impute as many missing GPS coordinates as possible based on available information. These GPS coordinates were missing mostly because of the weak GPS signals in indoor environments. We considered two scenarios. For Scenario A, there is missing GPS data during the night (23:00-8:00) with at least one hour of available GPS data. We imputed the missing GPS data by home addresses if the median GPS coordinates of the available GPS data during that night were within 100 m of home addresses [5]. Otherwise, we imputed the missing GPS directly by the median GPS coordinates during that night. For Scenario B, there is missing GPS data during the daytime (8:00-23:00). In this scenario, if the missing period was shorter than 6 hours and the distance between start and end positions of the missing period was shorter than 100 m, we imputed the missing GPS by the median GPS coordinates of the start and end positions. For missing GPS data that did not fall under these two scenarios, we did not impute due to limited information, which were subsequently dropped ($N$ = 936,687, 18.1%).

#### S2.2 Overview of context identification

We first applied the TrajDBSCAN algorithm, a clustering algorithm which extends the DBSCAN algorithm (details in **Section S2.3**) and can provide robust estimates of stay points – locations where an individual stays for a period of time – in sparse and noisy trajectories, to classify all data points into different clusters of stay points and trips [6-9]. The microenvironment of each cluster of stay points was determined by the median coordinates of this cluster. Land use data in 2010 were collected from the Washington

State Geospatial Portal [10], and were merged into seven types of land use, including commercial, industrial, residential, office, public facilities, park and open space, and other land use. The land use type of the nearest land use polygon was assigned to each cluster of stay points. We also obtained the building data from OpenStreetMap, and used a 10-m buffer of the building polygons to identify whether the stay points were located in indoor environments. To minimize the effect of measurement error of GPS coordinates, we did the following two post-processing steps: (1) Stay points which were during the hours of 23:00-8:00 and were within 100 m of home addresses were assigned as residential land use; (2) Stay points which covered 23:00-8:00 and were classified as residential were considered indoor. Since study recruitment occurred within Washington state, study participant travel outside of the state was rare, and the spatial land use data were obtained for Washington State, we excluded those stay points which were out of Washington State ($N$ = 235,718, 4.6%). Details for microenvironment classification of stay points are available in **Section S2.4**.

For data points detected as trips, we followed the method of Yi et al., and used the mean and standard deviation of speed and travel distance for each trip to classify all trips into vehicle-based trips and walking-based trips [5]. Detailed criteria for classifying trips are available in SM1 **Section S2.5**. All the vehicle-based trips were assigned as the vehicle microenvironment and considered as indoor activities, while the walking-based trips were assumed to be outdoor and assigned as the land use type of the nearest land use polygon. There was a total of 3,801,861 data points (73.7%) from 160 participants left after context identification.

#### S2.3 Identification of stay points

This study used the TrajDBSCAN algorithm to distinguish stay points from trips among all data points. The TrajDBSCAN algorithm is an improved clustering algorithm based on DBSCAN algorithm [6]. The pseudo code of the TrajDBSCAN algorithm is shown in **Figure S2** below. In this study, we selected the minimum time as 5 minutes and neighborhood maximum distance as 50 m.

**Figure S2.** Pseudo code of the TrajDBSCAN algorithm [6].

```
input  : Q //trajectory
         minTime //minimum time
         eps //neighborhood maximum distance
output: PS the set of personalized stops w.r.t minTime and eps
PS = ∅
foreach point Q_i in Q do
    if Q_i is unprocessed then
        mark Q_i as processed
        N = Eps-Linear-Neighbors(Q_i, eps)
        if duration(C) > minTime then
            C = ∅
            C = C ∪ Q_i
            foreach point Q_j in N do
                if Q_j is unprocessed then
                    C = C ∪ Q_j
                    N' = Eps-Linear-Neighbors(Q_j, eps)
                    if duration(N') > minTime then
                        N = N ∪ N'
            PS = PS ∪ C
return PS
```

#### S2.4 Microenvironment (ME) classification of stay points

The land use data in 2010 from the Washington State (WA) Geospatial Portal has 74 land use subtypes in total. In this study, we merged them into 7 major types, including commercial, industrial, residential, office, public facilities, park and open space, and other land use (**Figure S3**). The detailed criteria of this merging are shown in **Table S1** below. If the median coordinates of a cluster of stay points were right located in one land use polygon, the land use type of this polygon was directly assigned to these stay points (**Figure S4**). However, the WA Geospatial Portal database does not include roads, so if the median coordinates of a cluster of stay points were located on roads, we used the nearest land use polygon to classify the land use type for the stay points (**Figure S4**).

**Figure S3.** Land use and Building data.

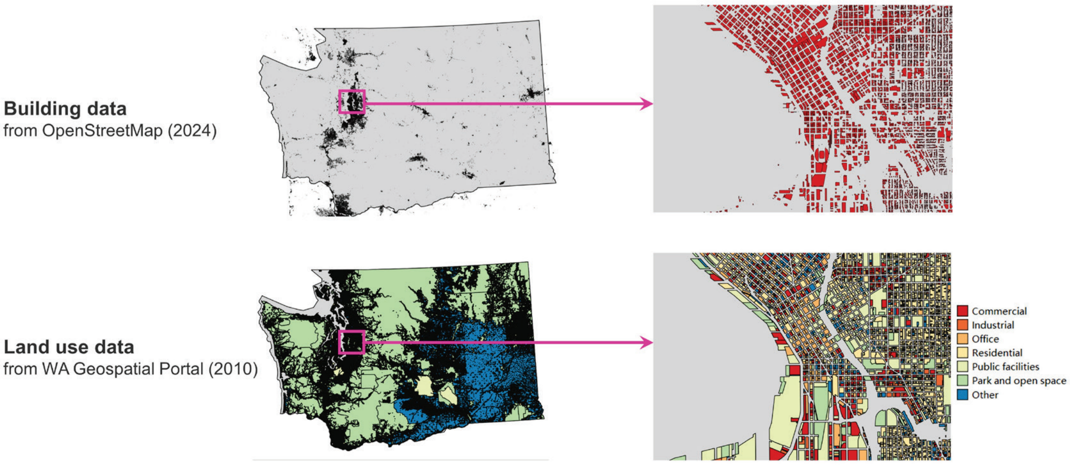


**Figure S4.** Schematics for ME classification.

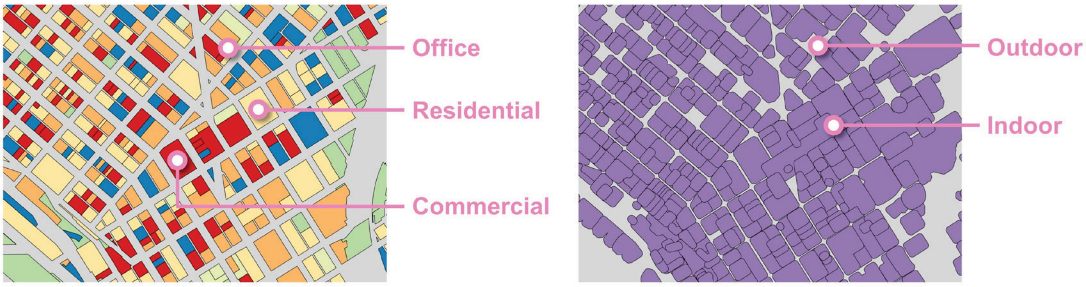


**Table S1.** The land use types used in WA Geospatial Portal and in this study.

| Code | Land use subtype in WA Geospatial Portal | Land use type in WA Geospatial Portal | Land use type in this study |
|---|---|---|---|
| 11 | Household, single-family units | Residential | Residential |
| 12 | Household, 2-4 units | Residential | Residential |
| 13 | Household, multi-units (5 or more) | Residential | Residential |
| 14 | Residential condominiums | Residential | Residential |
| 15 | Mobile home parks or courts | Residential | Residential |
| 16 | Hotels/motels | Residential | Residential |
| 17 | Institutional lodging | Residential | Residential |
| 18 | All other residential not elsewhere coded | Residential | Residential |
| 19 | Vacation cabin | Residential | Residential |
| 21 | Food and kindred products | Industrial | Industrial |
| 22 | Textile mill products | Industrial | Industrial |

| Code | Land use subtype in WA Geospatial Portal | Land use type in WA Geospatial Portal | Land use type in this study |
|---|---|---|---|
| 23 | Apparel and other finished products made from fabrics, leather, and simi | Industrial | Industrial |
| 24 | Lumber and wood products (except furniture) | Industrial | Industrial |
| 25 | Furniture and fixtures | Industrial | Industrial |
| 26 | Paper and allied products | Industrial | Industrial |
| 27 | Printing and publishing | Industrial | Industrial |
| 28 | Chemicals | Industrial | Industrial |
| 29 | Petroleum refining and related industries | Industrial | Industrial |
| 30 | Rubber and miscellaneous plastic products | Industrial | Industrial |
| 31 | Leather and leather products | Industrial | Industrial |
| 32 | Stone, clay and glass products | Industrial | Industrial |
| 33 | Primary metal industries | Industrial | Industrial |
| 34 | Fabricated metal products | Industrial | Industrial |
| 35 | Professional scientific, and controlling instruments, photographic | Industrial | Industrial |
| 39 | Miscellaneous manufacturing | Industrial | Industrial |
| 41 | Railroad/transit transportation | Transportation, communication, and utilities | Public facilities |
| 42 | Motor vehicle transportation | Transportation, communication, and utilities | NA |
| 43 | Aircraft transportation | Transportation, communication, and utilities | Public facilities |
| 44 | Marine craft transportation | Transportation, communication, and utilities | Public facilities |
| 45 | Highway and street right of way | Transportation, communication, and utilities | NA |
| 46 | Automobile parking | Transportation, communication, and utilities | Public facilities |
| 47 | Communication | Transportation, communication, and utilities | Public facilities |

| Code | Land use subtype in WA Geospatial Portal | Land use type in WA Geospatial Portal | Land use type in this study |
|---|---|---|---|
| 48 | Utilities | Transportation, communication, and utilities | Public facilities |
| 49 | Other transportation, communication, and utilities | Transportation, communication, and utilities | Public facilities |
| 51 | Wholesale trade | trade | Commercial |
| 52 | Retail trade - building materials, hardware, and farm equipment | trade | Commercial |
| 53 | Retail trade - general merchandise | trade | Commercial |
| 54 | Retail trade - food | trade | Commercial |
| 55 | Retail trade - automotive, marine craft, aircraft, and accessories | trade | Commercial |
| 56 | Retail trade - apparel and accessories | trade | Commercial |
| 57 | Retail trade - furniture, home furnishings and equipment | trade | Commercial |
| 58 | Retail trade - eating and drinking | trade | Commercial |
| 59 | Other retail trade | trade | Commercial |
| 61 | Finance, insurance, and real estate services | services | Office |
| 62 | Personal services | services | Office |
| 63 | Business services | services | Office |
| 64 | Repair services | services | Commercial |
| 65 | Professional services | services | Public facilities |
| 66 | Contract construction services | services | Commercial |
| 67 | Governmental services | services | Public facilities |
| 68 | Educational services | services | Public facilities |
| 69 | Miscellaneous services | services | Public facilities |
| 71 | Cultural activities and nature exhibitions | cultural, entertainment, and recreational | Public facilities |
| 72 | Public assembly | cultural, entertainment, and recreational | Park and open space |
| 73 | Amusements | cultural, entertainment, and recreational | Park and open space |
| 74 | Recreational activities | cultural, entertainment, and recreational | Park and open space |
| 75 | Resorts and group camps | cultural, entertainment, and recreational | Park and open space |
| 76 | Parks | cultural, entertainment, and recreational | Park and open space |

| Code | Land use subtype in WA Geospatial Portal | Land use type in WA Geospatial Portal | Land use type in this study |
|---|---|---|---|
| 79 | Other cultural, entertainment, recreational, church, cemetery | cultural, entertainment, and recreational | Public facilities |
| 81 | Agriculture (not classified under current use law) | Resource production and extraction | Other |
| 82 | Agriculture related activities | Resource production and extraction | Other |
| 83 | Agriculture classified under current use chapter 84.34 RCW | Resource production and extraction | Other |
| 84 | Fishing activities and related services | Resource production and extraction | Other |
| 85 | Mining activities and related services | Resource production and extraction | Other |
| 87 | Public timberland/non-designated forest | Resource production and extraction | Park and open space |
| 88 | Designated forest land under chapter 84.33 RCW | Resource production and extraction | Park and open space |
| 89 | Other resource production | Resource production and extraction | Other |
| 91 | Undeveloped land | Undeveloped land and water areas | Other |
| 92 | Noncommercial forest | Undeveloped land and water areas | Park and open space |
| 93 | Water areas | Undeveloped land and water areas | Park and open space |
| 94 | Open space land classified under chapter 84.34 RCW | Undeveloped land and water areas | Park and open space |
| 95 | Timberland classified under chapter 84.34 RCW | Undeveloped land and water areas | Park and open space |
| 99 | Other undeveloped land | Undeveloped land and water areas | Other |
| 100 | NA | | Other |

The building data were acquired from OpenStreetMap (**Figure S3**). We used a 10-m buffer of the building polygons to identify whether the stay points were located in indoor environments (**Figure S4**). The 10-m buffer was selected because we took the measurement error of GPS coordinates (approximately 15 m) and the average distance between building centroid and outline (approximately 5 m for all buildings across Washington State) into consideration. The measurement error of GPS coordinates was assessed by a pilot experiment. One PUWP monitor was placed in a house for about a day, where the true GPS coordinates are known. We calculated the distance between the coordinates from the PUWP and the true coordinates, and selected the 80$^{th}$ percentile as this measurement error (i.e., 15 m).

#### S2.5 Trip mode detection

We followed the procedures in Yi et al. to classify all trips into walking-based or vehicle-based trips [5], which is shown in **Figure S5** below. A threshold of 2 m/s (4.5 mph) was used to initially distinguish vehicle-based trips from walking-based trips. Since walking usually has a lower standard deviation (SD) of speed than low-speed driving, additional criteria about SD of speed and total distance of trips were added. We merged vehicle-based trips and likely vehicle-based trips into one type, vehicle-based trips, in this study, and did the same for walking-based trips.

**Figure S5.** Procedures of trip mode detection in this study.

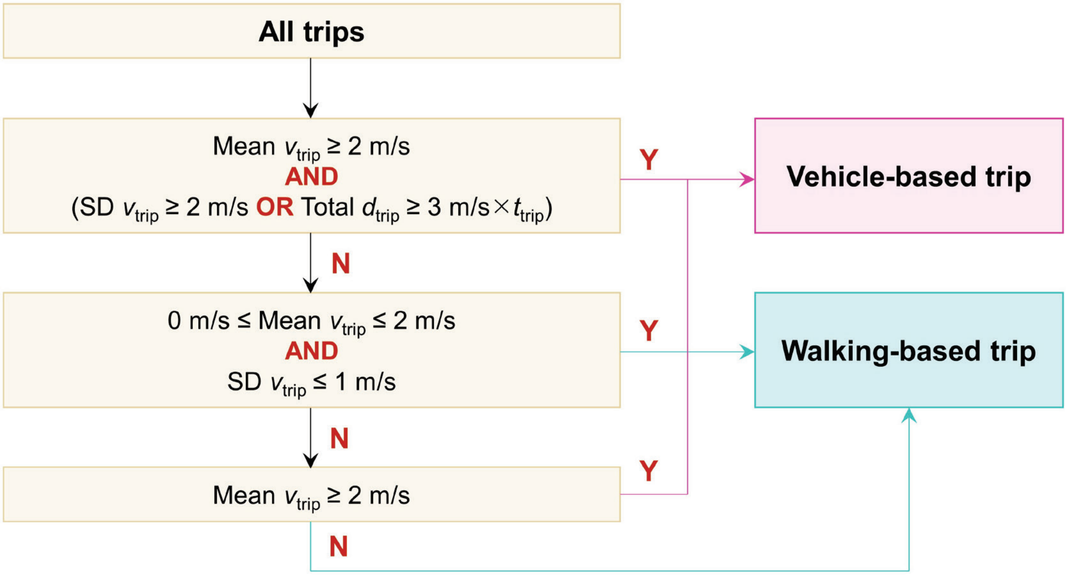

### Section S3 Calibration of air pollution data

#### S3.1 Methods of calibration

Before the formal calibration model, missing air pollution data were imputed. For each 1-minute particle measurement, if all particle counts for individual size bins were zero, the PNC of this data point was imputed with the average PNC of the 5 nearest time points, which was approximately the 5-min average level. We dropped the data points which could not be imputed due to absence of valid temporally-near PNC data ($N$ = 66,127, 1.3%).

We employed a 2-step calibration modeling approach, in which the first step aimed to model the sensor's relationship between size-specific PNC measurements and the sensor manufacturer's $PM_{2.5}$ measurement, and the second step aimed to model the relationship between participants' $PM_{2.5}$ sensor outdoor measurements and nearby $PM_{2.5}$ measurements collected by reference ambient air quality monitoring sites. The reason why we selected a 2-step calibration approach is to take full advantage of available data to obtain a calibration model that is as generalizable as possible. First, due to the rounding errors of $PM_{2.5}$ mass concentrations to integers from the PUWP, directly using the $PM_{2.5}$ mass concentration readings was not so accurate, especially for low exposures. About 40% of data points have positive PNC measurements with low count numbers but zero $PM_{2.5}$ mass concentrations, which was not possible in reality. Second, although the second step could only use outdoor hourly-level measurement data, the first step can include all indoor and outdoor minute-level data into the model. Third, while the ambient air quality data used in the second step tend to be within a relatively low concentration range, the first step allows a wide range of concentrations to be considered.

In the first modeling step, the sensor's $PM_{2.5}$ measurement at 1-min interval is related to the sensor's PNC measurements in different size bins, with quadratic terms included for possible non-linear temperature and humidity effects on the relationship [11, 12].

$$PM_{2.5m,sensor} \sim T + T^2 + RH + RH^2 + PM_{0.3c} + PM_{0.5c} + PM_{1c} + PM_{2.5c} + PM_{5c} \quad (S1)$$

where $PM_{2.5m,sensor}$ is the $PM_{2.5}$ mass concentration provided by the sensor, μg/m$^3$; $T$ is the air temperature, °C; $RH$ is the relative humidity, ranging from 0 to 100; $PM_{0.3c}$, $PM_{0.5c}$, $PM_{1c}$, $PM_{2.5c}$, and $PM_{5c}$ are PNC in five size bins (i.e., >0.3 μm, >0.5 μm, >1 μm, >2.5 μm, and >5 μm), respectively, counts/100mL. The inclusion criteria of data points for this regression model included: (1) The sensor data should satisfy the inequalities, i.e., $PM_{0.3c} \geq PM_{0.5c} \geq PM_{1c} \geq PM_{2.5c} \geq PM_{5c}$ and $PM_{1m,sensor} \leq PM_{2.5m,sensor} \leq PM_{10m,sensor}$; (2) The temperature is lower than 40°C to exclude a possible abnormal condition for the sensor; (3) The $PM_{2.5}$ mass concentration is lower than 229 μg/m$^3$ to exclude abnormally high concentrations. (Note: This threshold, 229 μg/m$^3$, is the 10$^{th}$ percentile of $PM_{2.5}$ mass concentration when $PM_{0.3c}$ overflows to negative values, suggesting a too high particle number concentration (> 32767 counts/100mL).) Then the prediction of this first-step calibration model can be obtained and denoted as $\widehat{PM_{2.5m,sensor}}$. We constrained $\widehat{PM_{2.5m,sensor}}$ equal to zero if the prediction value was negative (accounting for 0.35%).

The second step used an in-situ calibration approach to compare the sensor data when the microenvironment context identifies that the sensor is outdoors with the monitoring data from regulatory monitoring stations. We assumed that the outdoor $PM_{2.5}$ mass concentration could be estimated using the nearest regulatory monitoring data if the nearest regulatory monitoring station was relatively close to the outdoor sensor location. Therefore, we selected the eligible data points through the following criteria: (1) The data points were outdoor stay points or walking-based trips; (2) The sensor data should satisfy the inequality, i.e., $PM_{0.3c} \geq PM_{0.5c} \geq PM_{1c} \geq PM_{2.5c} \geq PM_{5c}$; (3) The temperature is lower than 40°C; (4) All PNCs should be lower than 95$^{th}$ percentile of the corresponding size bin to avoid the effect of extreme values; and most importantly, (5) The GPS coordinates should be within the cut-off distance from the regulatory monitoring stations in the Washington Air Monitoring Network [13]. In this study, we tried different cut-off distances, including 0.5, 0.6, 0.7, 0.8, 0.9, 1, 2, 3, and 5 km, to maximize the goodness-of-fit of the calibration model. As the regulatory monitoring stations provide hourly average $PM_{2.5}$ concentrations, we also aggregated the eligible minute-level sensor data to an hourly average level to match the time scale of data. We finally included the hourly average data points, which contained at least 20 eligible minute-level sensor data in one hour to guarantee the accuracy of estimating the hourly average, into the second-step calibration. A linear mixed model with random slope was applied for the second-step calibration, as shown in equation (S2).

$$PM_{2.5m,nearest} \sim \widehat{PM_{2.5m,sensor}} + \left(\widehat{PM_{2.5m,sensor}} \mid studyid\right) \quad \text{(S2)}$$

where $PM_{2.5m,nearest}$ is the hourly average $PM_{2.5}$ concentration from the nearest regulatory monitoring station, μg/m$^3$; $\widehat{PM_{2.5m,sensor}}$ is the hourly average of the first step minute-level prediction; *studyid* is the identification number of each participant. Then, the calibrated $PM_{2.5}$ mass concentration (denoted as $PM_{2.5m,calibrated}$) could be predicted by the above model, and negative predictions were considered as zeros. If the sample size was too limited to support a random slope model, a random intercept model was used. The data points that did not satisfy the inequality, i.e., $PM_{0.3c} \geq PM_{0.5c} \geq PM_{1c} \geq PM_{2.5c} \geq PM_{5c}$, were excluded in this prediction ($N = 23{,}509$, 0.5%).

#### S3.2 Results of calibration

**Figure S6*a*** shows the performance of the first-step calibration model. After considering the effect of temperature and relative humidity, the predicted $PM_{2.5}$ concentration of this model agrees well with the observed $PM_{2.5}$ concentration directly provided by the PUWP sensors over a wide range of mass concentrations ($R^2$ = 0.97, RMSE = 3.0 μg/m$^3$). Detailed parameters of the first-step calibration model are available in **Table S2**. For the second-step calibration model, the model performance under different cut-off distances is shown in **Table S3**. The $R^2$ of the calibration model dropped from 0.98 to 0.56 when the cut-off distance increased from 0.5 km to 5 km. We considered 0.8 as a threshold of $R^2$, and attempted to include as many data points as possible into the second-step calibration model, so we finally selected 0.6 km as the cut-off distance for this study. As is shown in **Figure S6*b***, the calibrated $PM_{2.5}$ hourly average concentrations are highly correlated with those from the nearest regulatory monitoring stations ($R^2$ = 0.93, RMSE = 0.1 μg/m$^3$), suggesting the high performance of this in-situ calibration. Detailed parameters of the second-step calibration model are shown in **Table S4**. The $PM_{2.5}$ concentrations from the sensors before calibration are approximately 30% higher than those from the regulatory monitoring stations. This is consistent with previous studies in that the light-scattering $PM_{2.5}$ sensors usually overestimate the $PM_{2.5}$ mass concentrations, compared to gravimetric instruments [14-17].

**Figure S6.** Results of the calibration models: (*a*) Relationship between predicted $PM_{2.5}$ concentration of the first-step calibration model and observed $PM_{2.5}$ concentration of sensors; (*b*) Relationship between $PM_{2.5}$ concentration of sensors within 600 m of regulatory monitoring stations calibrated by the second-step calibration model and $PM_{2.5}$ concentration from corresponding regulatory monitoring stations.

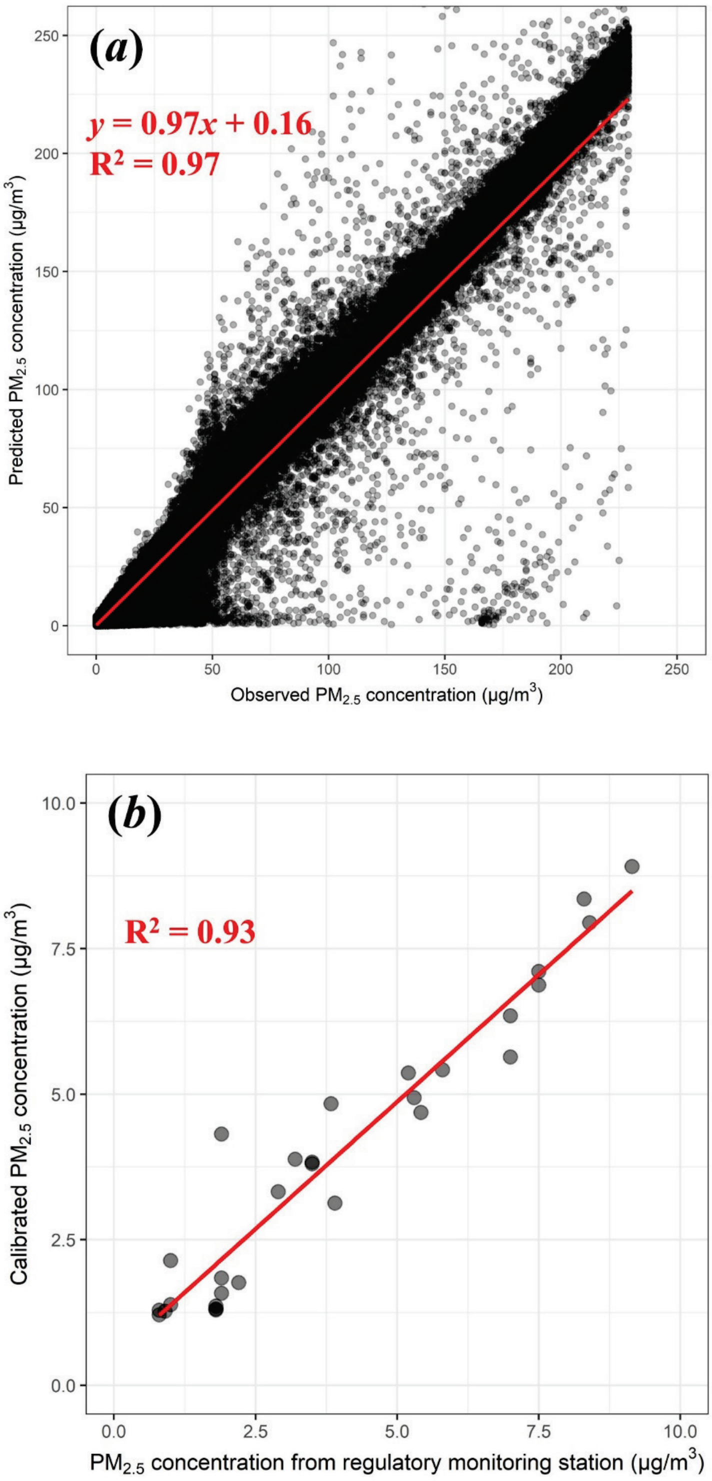

**Table S2.** Parameters of first-step calibration model.

| Covariate | Coefficient | SE | *p*-value |
|---|---|---|---|
| (Intercept) | -0.334 | 0.0521 | <0.001 |
| RH | 0.0551 | 0.00142 | <0.001 |
| $RH^2$ | $-5.33\times10^{-4}$ | $1.95\times10^{-5}$ | <0.001 |
| T | -0.0723 | 0.00368 | <0.001 |
| $T^2$ | 0.00183 | $7.02\times10^{-5}$ | <0.001 |
| $PM_{0.3c}$ | -0.00127 | $5.80\times10^{-6}$ | <0.001 |
| $PM_{0.5c}$ | 0.0196 | $2.22\times10^{-5}$ | <0.001 |
| $PM_{1c}$ | 0.0344 | $3.15\times10^{-5}$ | <0.001 |
| $PM_{2.5c}$ | 0.0409 | $2.64\times10^{-4}$ | <0.001 |
| $PM_{5c}$ | -0.0648 | $4.98\times10^{-4}$ | <0.001 |

**Table S3.** Relationship between the cut-off distance and $R^2$ of the second-step calibration model.

| Cut-off Distance (km) | Concentration range (μg/m$^3$) | Sample size | $R^2$ | RMSE (μg/m$^3$) | Slope |
|---|---|---|---|---|---|
| 0.5* | 1.8 – 9.2 | 19 | 0.979 | 0.038 | 0.502 |
| 0.6 | 0.8 – 9.2 | 29 | 0.926 | 0.084 | 0.785 |
| 0.7 | 0.8 – 11.6 | 57 | 0.780 | 0.161 | 0.558 |
| 0.8 | 0.8 – 21.0 | 151 | 0.709 | 0.198 | 0.366 |
| 0.9 | 0.8 – 21.0 | 201 | 0.629 | 0.273 | 0.240 |
| 1 | 0.8 – 21.0 | 228 | 0.672 | 0.264 | 0.378 |
| 2 | 0.0 – 27.5 | 906 | 0.455 | 0.294 | 0.317 |
| 3 | 0.0 – 42.5 | 1557 | 0.543 | 0.356 | 0.513 |
| 5 | 0.0 – 42.5 | 3150 | 0.561 | 0.347 | 0.548 |

* Random intercept model was applied due to too limited sample size. Other scenarios used random slope model.

**Table S4.** Parameters of second-step calibration model.

| Covariate | Coefficient | SE | *p*-value |
|---|---|---|---|
| *Fixed effects* | | | |
| (Intercept) | 3.346 | 0.959 | 0.007 |
| $PM_{2.5m,sensor}$ | 0.785 | 0.395 | 0.095 |
| *Random effects* | | | |
| (Intercept) | 5.888 | 2.427 | Corr |
| $PM_{2.5m,sensor}$ | 0.783 | 0.885 | -0.84 |

**Figure S7.** Scatterplots for second-step calibration model with different cut-off distances.

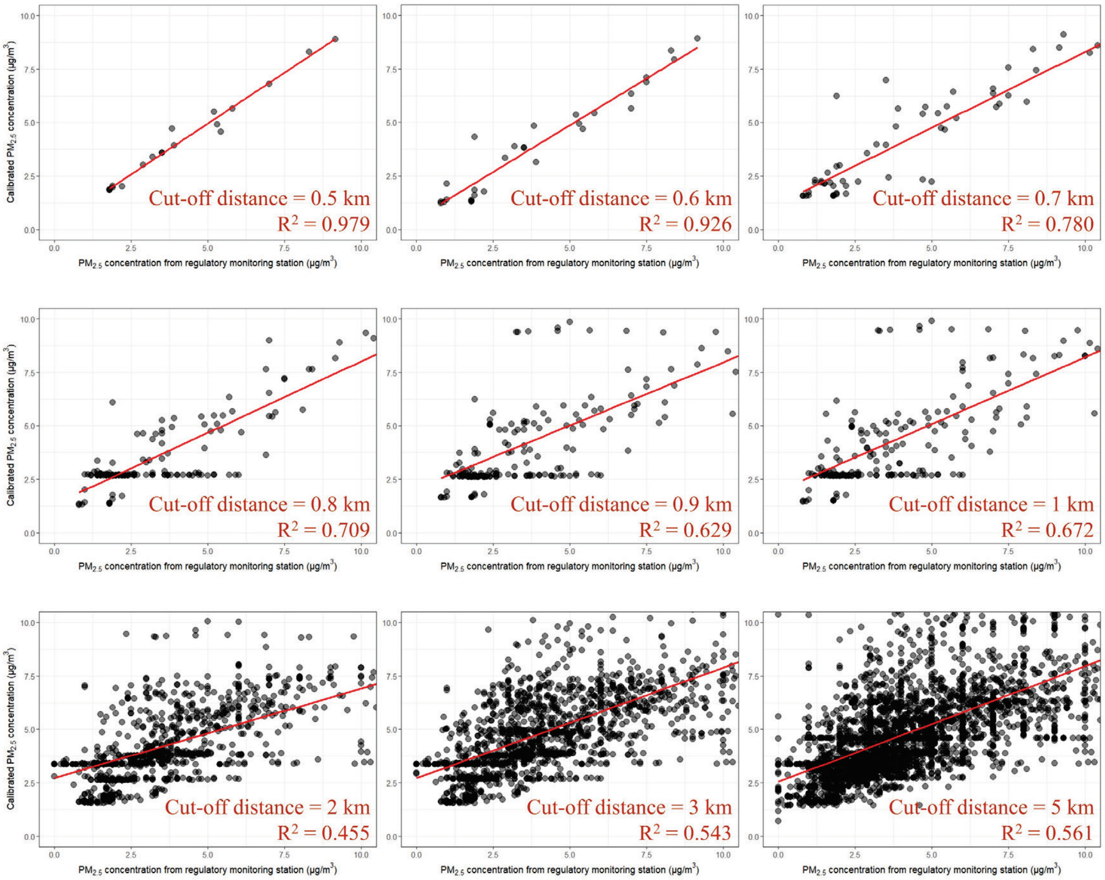

### Section S4 Exposure assessment

#### S4.1 Methods of exposure assessment

Although data for this study were collected from twin pairs, we assessed individual-level mean hourly $PM_{2.5}$ exposures during the two-week monitoring period. This part of the analysis did not rely on the microenvironment identification, so we used as many eligible data points as possible (i.e., methods described in Sections 2.3.1 and 2.3.3). A total of 4,874,563 data points (94.4%) from 163 participants were included. Personal exposure concentrations of $PM_{2.5}$ were compared among different demographic and socioeconomic status (SES) characteristics, including age, sex, race, marital status, highest education level, and annual household income. Age groups were divided into 0-29, 30-39, 40-49, 50-59, and ≥60 years old. Sex groups were male and female. Race was classified as white and non-white. Marital status was divided into currently married and unmarried. The highest education level was classified into three types, including lower than a bachelor degree (BA), BA, and higher than BA. We further considered low and high annual household income groups, which referred to $90,000, the median in Washington State, as a threshold.

Next, we assessed the spatiotemporal patterns of the personal exposures based on microenvironment context identification (methods in Section 2.3.2), with a total of 3,712,225 data points (71.9%) from 160 participants were included. In order to obtain the more accurate time spent in different microenvironments and contribution of each microenvironment to the total exposure dose, we excluded the invalid participant-days which had less than 6 hours for one participant [5]. From the spatial perspective, personal exposure concentrations of $PM_{2.5}$ in eight microenvironments (including seven land use types and vehicles) and indoor/outdoor environments were summarized across the valid days in the two-week monitoring period and compared with each other. To evaluate the contribution of each microenvironment to the cumulative exposure, the proportion of exposure dose for the *k*th microenvironment (denoted as $Proportion_k$) was further calculated by equation (S3) below.

$$\text{Proportion}_k = \frac{\text{Exposure dose}_k}{\text{Exposure dose}_{total}} = \frac{\sum_i \text{PM}_{2.5\text{m,calibrated}(i)} \delta_{ik} Q \Delta t_i}{\sum_i \text{PM}_{2.5\text{m,calibrated}(i)} Q \Delta t_i} \qquad \text{(S3)}$$

where $PM_{2.5\text{m,calibrated}(i)}$ is the calibrated $PM_{2.5}$ mass concentrations at time $i$, µg/m$^3$; $\triangle t_i$ is the sampling interval at time $i$, second; $\delta_{ik}$ is 1 if the exposure occurs in the $k^{th}$ microenvironment at time $i$ and 0 otherwise; $Q$ is the inhalation rate which was assumed constant and will then be canceled out in both the numerator and denominator, m$^3$/s. From the temporal perspective, we compared the personal $PM_{2.5}$ exposure concentrations in different seasons (winter: December to February, spring: March to May, summer: June to August, autumn: September to November) and hours in a day.

In addition, for comparisons with the personal exposure monitoring data, we assessed the hourly average exposure concentrations at participants' residential location based only on regulatory monitoring station data, which is an approach used in many epidemiological studies. We applied the inverse distance weighted (IDW) interpolation to obtain the outdoor hourly average $PM_{2.5}$ concentrations at participants' home addresses from the regulatory monitoring network data [18-22]. In detail, we used the inverse of the squared distance from home addresses to each regulatory monitoring station in WA as the weight function for the IDW interpolation. Then we compared the exposure concentrations from the home-based approach with those GPS-based personal monitoring results.

#### S4.2 Results of exposure assessment

**Figure S8.** Boxplots of (*a*) medians and (*b*) 97.5th percentiles of participant-level personal $PM_{2.5}$ exposure with different demographic and SES characteristics. (* $p < 0.05$)

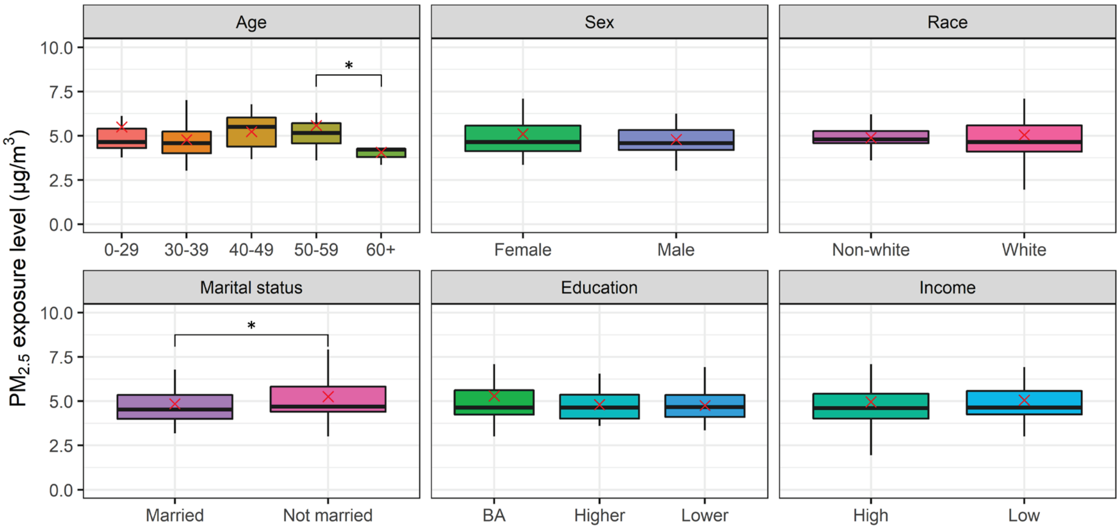


(*a*)

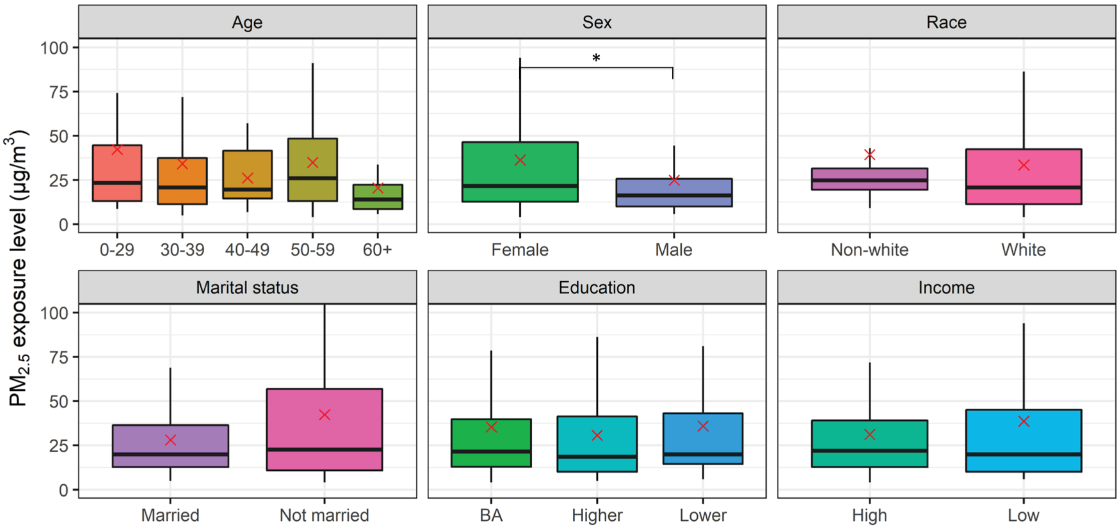


(*b*)

**Figure S9.** Personal $PM_{2.5}$ exposure concentrations with GPS coordinates for participant AIR4585A in two days in April, 2018.

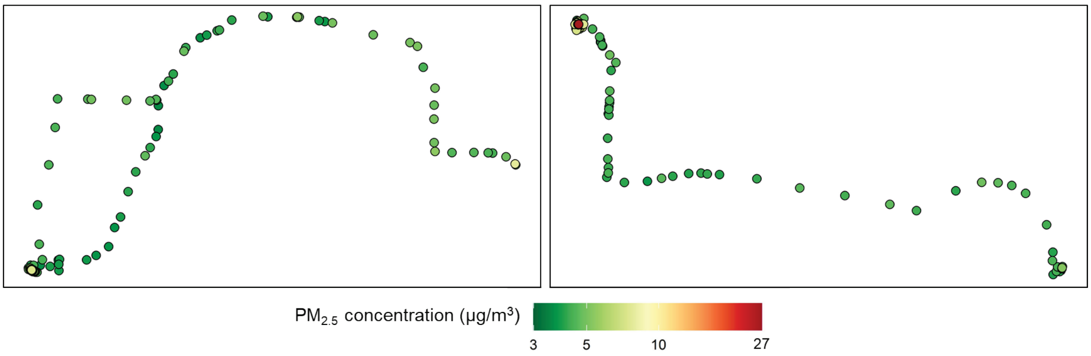


**Figure S10.** Time fraction spent in indoor/outdoor environments and different types of land use.

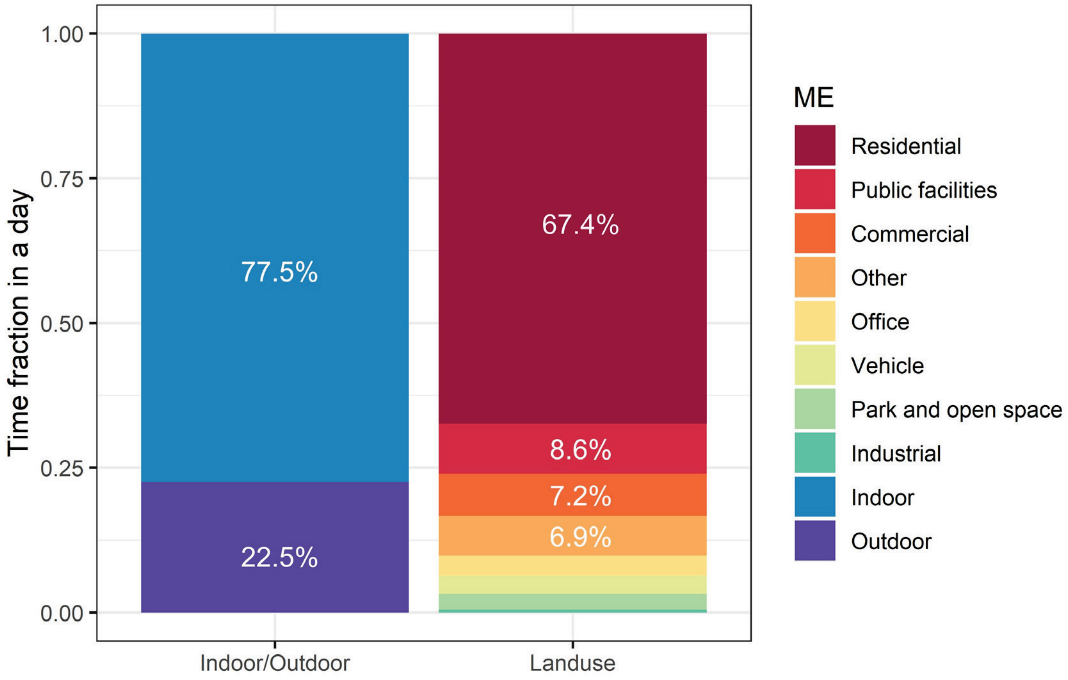

**Figure S11.** Comparison between home-based assessment and GPS-based personal monitoring results in this study for (*a*) AIR4585A in April, 2018, and (*b*) AIR4602A in August, 2018. Note: Grey time period refers to outdoor activities.

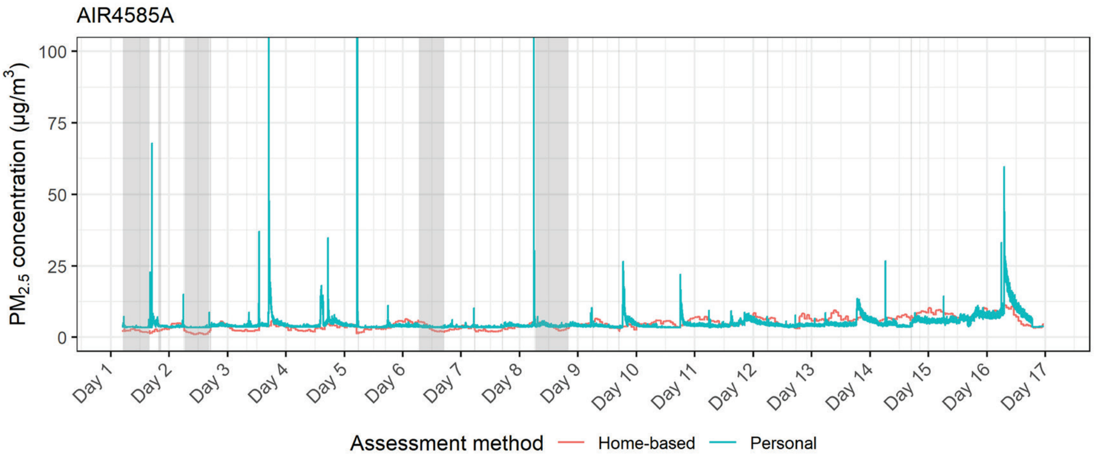


(*a*)

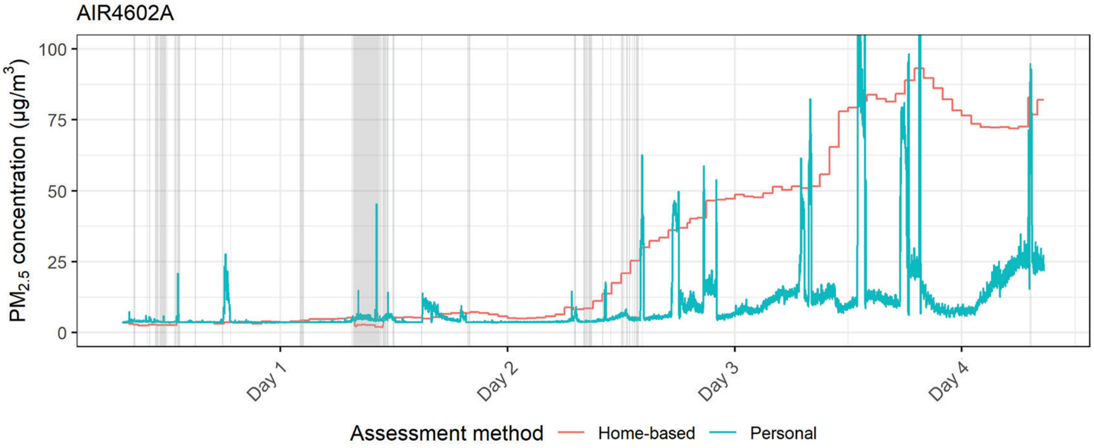


(*b*)

#### Section S5 Comparison between within-participant and between-participant variations

A variance component analysis was performed using a random intercept model, which is shown below.

$$\mathrm{PM}_{2.5\mathrm{m,calibrated}} \sim 1+\left(1\middle|studyid\right) \quad \text{(S4)}$$

where $\mathrm{PM}_{2.5\mathrm{m,calibrated}}$ is the calibrated mass concentration of personal $\mathrm{PM}_{2.5}$ exposure concentration; and *studyid* is the identity number of each participant. The intraclass correlation coefficient (ICC) was calculated as

$$ICC = \frac{\tau_0^2}{\tau_0^2 + \sigma^2} \quad \text{(S5)}$$

where $\tau_0{}^2$ is the interclass variance, and $\sigma^2$ is the intraclass variance. A lower ICC represents a larger variation within each participant compared to the variation between participants.

12. Zusman, M.; Schumacher, C. S.; Gassett, A. J.; Spalt, E. W.; Austin, E.; Larson, T.

V.; Carvlin, G.; Seto, E.; Kaufman, J. D.; Sheppard, L., Calibration of low-cost particulate matter sensors: Model development for a multi-city epidemiological study. *Environ Int* **2020,** *134*, 105329.

13. Department of Ecology State of Washington Washington Air Quality Map. https://enviwa.ecology.wa.gov/mobile/ (Aug 12, 2024),

14. Shi, J. J.; Chen, F. E.; Cai, Y. F.; Fan, S. C.; Cai, J.; Chen, R. J.; Kan, H. D.; Lu, Y. H.; Zhao, Z. H., Validation of a light-scattering PM2.5 sensor monitor based on the long-term gravimetric measurements in field tests. *Plos One* **2017,** *12*, (11), e0185700.

15. McNamara, M. L.; Noonan, C. W.; Ward, T. J., Correction Factor for Continuous Monitoring of Wood Smoke Fine Particulate Matter. *Aerosol Air Qual Res* **2011,** *11*, (3), 316-323.

16. Li, Z. Y.; Che, W. W.; Lau, A. K. H.; Fung, J. C. H.; Lin, C. Q.; Lu, X. C., A feasible experimental framework for field calibration of portable light-scattering aerosol monitors: Case of TSI DustTrak. *Environ Pollut* **2019,** *255*, 113136.

17. Moazami, T. N.; Svendsen, K. V.; Buhagen, M.; Jorgensen, R. B., Comparing PM2.5, respirable dust, and total dust fractions using real-time and gravimetric samples in an exposure chamber study. *Heliyon* **2023,** *9*, (6), e16127.

18. Deng, Q. H.; Lu, C.; Norback, D.; Bornehag, C. G.; Zhang, Y. P.; Liu, W. W.; Yuan, H.; Sundell, J., Early life exposure to ambient air pollution and childhood asthma in China. *Environ Res* **2015,** *143*, 83-92.

19. Pereira, G.; Bracken, M. B.; Bell, M. L., Particulate air pollution, fetal growth and gestational length: The influence of residential mobility in pregnancy. *Environ Res* **2016,** *147*, 269-274.

20. Shukla, K.; Kumar, P.; Mann, G. S.; Khare, M., Mapping spatial distribution of particulate matter using Kriging and Inverse Distance Weighting at supersites of megacity Delhi. *Sustain Cities Soc* **2020,** *54*, 101997.

21. Jung, J. Y.; Park, J. Y.; Kim, Y. C.; Lee, H.; Kim, E.; Kim, Y. S.; Lee, J. P.; Kim, H., Effects of air pollution on mortality of patients with chronic kidney disease: A large observational cohort study. *Sci Total Environ* **2021,** *786*, 147471.

22. Power, M. C.; Bennett, E. E.; Lynch, K. M.; Stewart, J. D.; Xu, X.; Park, E. S.; Smith, R. L.; Vizuete, W.; Margolis, H. G.; Casanova, R.; Wallace, R.; Sheppard, L.; Ying, Q.; Serre, M. L.; Szpiro, A. A.; Chen, J. C.; Liao, D.; Wellenius, G. A.; van Donkelaar, A.; Yanosky, J. D.; Whitsel, E., Comparison of PM2.5 Air Pollution Exposures and Health Effects Associations Using 11 Different Modeling Approaches in the Women's Health Initiative Memory Study (WHIMS). *Environ Health Perspect* **2024,** *132*, (1), 17003.
